# Polygenic effects on cortical size and ADHD converge on a mid-gestational progenitor-to-neuron transition programme

**DOI:** 10.64898/2026.09.21.26363544

**Authors:** Yuankai He, Amir Ebneabbasi, Koen Rademaker, Elina Jin, Yuanjun Gu, Fani Femi, Clara M.L. Riegis, Bess Pearson, Renato Polimanti, Jakob Grove, Anders D. Børglum, Nadine Parker, Ole Andreassen, Oleksandr Frei, Darren Cameron, Nicholas Bray, Ang Li, Jian Zeng, Naomi Wray, Rafael Romero-Garcia, Timothy Rittman, Simon Baron-Cohen, Melissa Gladstone, Shivaram Avula, Mary-Ellen Lynall, Sarah Rae, Duncan Astle, Richard A.I. Bethlehem, Edward T. Bullmore, Omer Ali Bayraktar, Varun Warrier

## Abstract

Neurodevelopmental and psychiatric conditions have long been hypothesised to emerge from changes in the structure and function of the brain. However, the molecular and cellular mechanisms linking brain structure to these conditions remain unknown. Here, using genetic and structural/diffusion neuroimaging data from 59,283 individuals spanning adults, children and neonates, we identify widespread genetic associations between ADHD, anxiety and depression and multiple phenotypes indexing cortical size and subcortical volume. Multivariate genetic analyses demonstrate that these associations are largely driven by a common cortical size genetic factor and ADHD. Mendelian randomisation indicates that genetically predicted reduced cortical size is causal for ADHD and not *vice versa*. Using single-nucleus RNA sequencing of the developing cortex, we further demonstrate that although cortical size and ADHD are enriched in different cell types - proliferating progenitors and postmitotic neurons, respectively, they converge on neurogenic genetic programmes active during the transition between the two cell types. This programme peaks at mid-gestation and is localised to the outer subventricular and intermediate zones, the regions where progenitor amplification and early neuronal migration drive human cortical expansion, and is enriched for genes that are differentially expressed in human and macaque developing cortex. Separately, the negative genetic correlation between ADHD and cortical size measures are strongest in cortical regions that have expanded most in humans relative to macaques. Together, these findings identify a novel, spatiotemporally restricted molecular mechanism underlying neurogenesis that is enriched for polygenic effects of both cortical size and ADHD.

## Introduction

Psychiatric and neurodevelopmental conditions (henceforth, neuropsychiatric conditions) have long been hypothesised to arise from changes to the structure and function of the human brain. Over two decades of large-scale Magnetic Resonance Imaging (MRI) have sought to identify these differences. These include, for example, reduced cortical surface area and grey matter volume in attention-deficit hyperactivity disorder (ADHD) and schizophrenia (*1* –*4* ); larger ventricles and smaller hippocampi in schizophrenia (*5* ). Smaller, and less robust differences have been reported in other neuropsychiatric conditions as well (*6* ). This body of work has identified candidate brain regions associated with neuropsychiatric conditions. Nevertheless, linking this to underlying biological mechanisms has been challenging for two reasons.

First, neuroimaging alone cannot establish whether these differences are causal for neuropsychiatric conditions, arising from shared genetic or non-genetic factors, or are sequelae of the conditions themselves. In schizophrenia, for example, accounting for illness duration and medication use attenuates volumetric differences (*4, 7* ). Large-scale datasets combining neuroimaging and genetics offer one way to resolve this. Genotype is fixed at conception, and liability to these conditions is continuously distributed in the population. Correlations between genetic liability and neuroimaging measures in unaffected cohorts are therefore unlikely to be driven by the consequences of illness or its treatment. A few studies have taken this approach, but have focused on one condition at a time (*8* –*10* ), or a limited set of MRI measures (*11* –*13* ), or been conducted in modestly sized cohorts leading to inconsistent results across studies (*14* ). In addition to inconsistent results, we also do not know how widely shared these genetic effects are across neuropsychiatric conditions and MRI measures, or when in development they emerge.

Second, neuroimaging alone cannot identify the molecular and cellular mechanisms through which brain structure and neuropsychiatric risk are linked. The structure of the human brain emerges through a series of coordinated, often transient, molecular and cellular processes that begin in the first trimester and continue into adolescence and beyond (*15* ). Any genetic effect shared between brain structure and neuropsychiatric risk must converge on one or more of these processes. But MRI alone, which typically measures tissue properties at the millimetre scale, cannot identify these processes. Consequently, we do not know in which cells and molecular mechanisms, and at what point in development this shared genetics converges. Single-nucleus and spatial transcriptomic atlases of the human brain now make these questions tractable.

Here, combining neuroimaging, genomic and transcriptomic datasets, we address both of these gaps: transdiagnostic causality and cellular neurodevelopmental mechanisms. First, using genetic correlations and polygenic scores, we assessed the shared genetics between six neuropsychiatric conditions (ADHD, autism, anxiety, depression, bipolar disorder, and schizophrenia) and 2,326 MRI-derived phenotypes (IDPs) spanning cortical and subcortical structure and white-matter microstructure in adults (UK Biobank, *N*_max_ = 53,709), children (ABCD, *N*_max_ = 5,119) and neonates (dHCP, N = 455) (*16* –*19* ). We then used multivariate genetic modelling (*20* ) and Mendelian randomisation (*21* ) to test whether these associations reflect a shared latent factor, and whether the genetically predicted effects are causal. To identify underlying mechanisms, we mapped the convergence of polygenic effects on brain structure and neuropsychiatric risk onto cell types, gene programmes, anatomical locations and evolutionary molecular signatures in the developing brain, using single-nucleus RNA sequencing (snRNA-seq) and spatial transcriptomic data.

### Widespread negative genetic correlations between brain size and ADHD, depression, and anxiety

Using linkage disequilibrium score regression (LDSC) (*22, 23* ), we first quantified genetic correlations between six well-powered neuropsychiatric conditions - ADHD, autism, anxiety, major depressive disorder (MDD), bipolar disorder, and schizophrenia (**Table S1**) - and 2,326 structural and diffusion-based IDPs measured in 53,709 individuals from the UK Biobank (*24* ). These included cortical, subcortical, ventricular, cerebellar, brain stem, and corpus-callosal volumes; measures of cortical size, curvature, and thickness; and five diffusion tensor imaging-derived measures of microstructural integrity assessed cortically and in white-matter tracts (**Box 1**). Cortical phenotypes were generated both globally (summed or averaged across the cortex) and across 180 bilaterally averaged regions defined by the Glasser parcellation (*25* ).

**BOX 1: IDPs investigated in this study**

We analysed 2,326 IDPs: 12 global measures, 2,159 regional cortical IDPs (12 phenotypes *×* 180 bilaterally averaged Glasser parcels, excluding one IDP of low variance), 20 subcortical, ventricular and corpus callosal volumes, and 135 white-matter tract IDPs.

**Macrostructural measures (T1-weighted MRI)** capture the size and shape of the cortex. Surface area (SA), grey matter volume (GMV) and the two folding measures (intrinsic curvature index (IC) and folding index (FI)) together form the cortical size factor given their high genetic similarity. Cortical thickness (CT) instead reflects the radial dimension of the cortex, and is genetically largely distinct from cortical size. Mean and Gaussian curvature (MC, GC) capture local cortical shape. We additionally measured subcortical, ventricular, cerebellar, brain stem, and corpus callosal volumes.

**Microstructural measures (diffusion-weighted MRI):** Fractional anisotropy (FA) and mean diffusivity (MD) index the directionality and overall magnitude of water diffusion, and are sensitive to membrane density, myelination and axonal organisation. Neurite Orientation Dispersion and Density Imaging additionally quantifies neurite density (ICVF), the degree to which neurites are aligned or dispersed (OD), and the free-water fraction (ISOVF).

We identified moderate negative genetic correlations between ADHD and IDPs related to brain size (cortical and subcortical volumes, surface area, folding index, and intrinsic curvature) at global level (*r*_*g*_ =-0.131 to -0.180), and with 91 regional phenotypes primarily in frontotemporal regions at a Bonferroni-corrected significance threshold of *p <* 8.12 *×* 10^−6^ (**Figure 1a, Table S2, Methods**). Additionally, we identified modest negative genetic correlation between the same IDPs and anxiety and MDD (*r*_*g*_ =-0.107 to -0.167, **Figure S1**).

**Figure 1:**
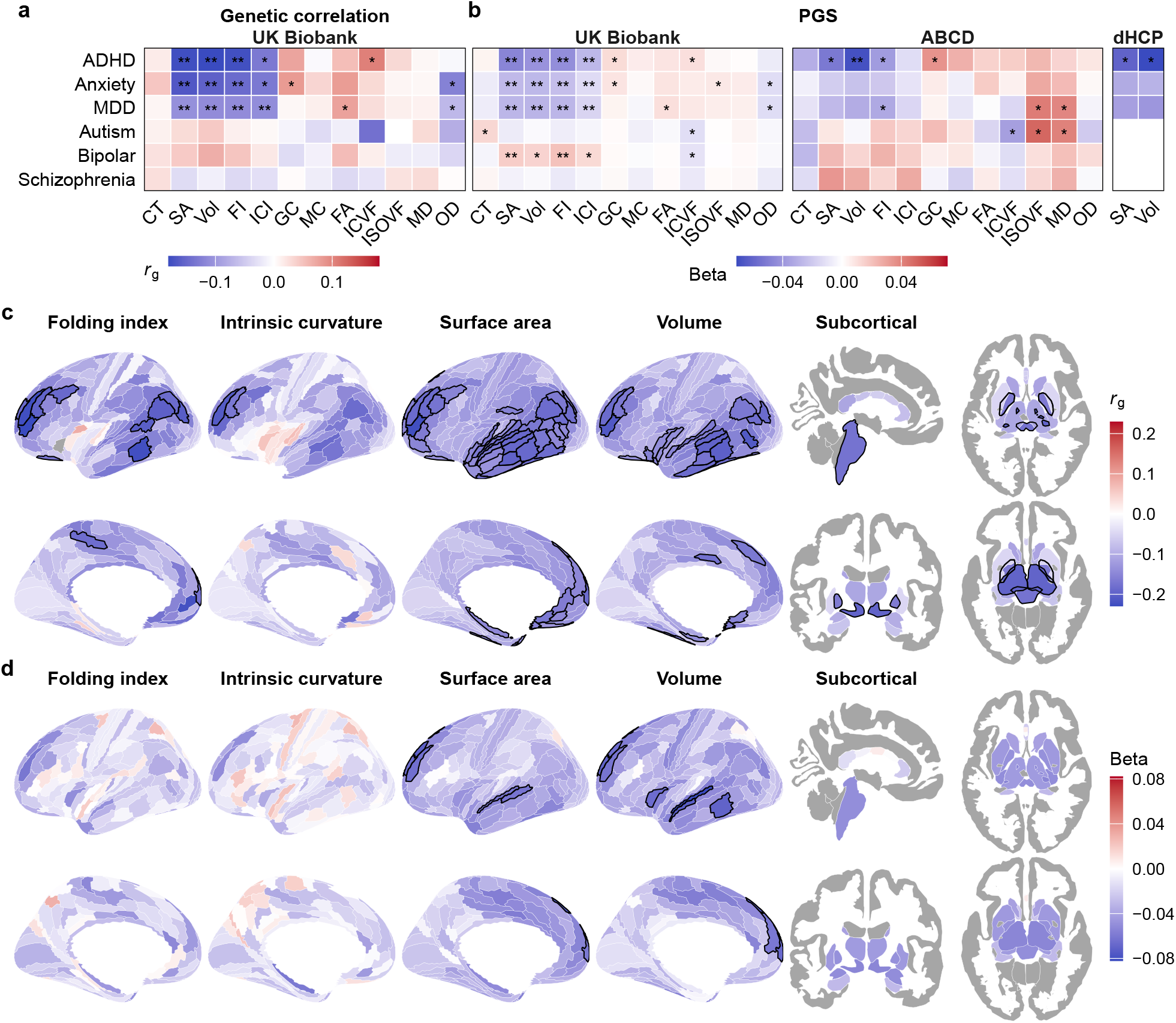
Genome-wide correlations between imaging and psychiatric phenotypes. **a** Genetic correlations (*r*_*g*_) between six neuropsychiatric conditions and 12 global imaging phenotypes, UK Biobank (*N* = 53, 709). **b** Effect sizes of polygenic scores of the same conditions on global imaging phenotypes in UK Biobank (*N* = 53, 709), ABCD (*N* = 5, 119) and dHCP (*N* = 455; surface area and volume only). **c** Regional *r*_*g*_ between ADHD and four cortical size measures across 180 Glasser parcels plus subcortical volumes, UK Biobank. **d** Polygenic score effect sizes of ADHD on regional cortical size measures across 180 Glasser parcels plus subcortical volumes, estimated in ABCD. **: *p <* 8.12 *×* 10^*-*6^ (Bonferroni); *: *q <* 0.05 (BH within condition). Outlined regions in **c**-**d** pass Bonferroni. Left hemisphere only (bilateral parcellation). CT: cortical thickness; SA: surface area; Vol: volume; FI: folding index; IC: intrinsic curvature; GC: Gaussian curvature; MC: mean curvature; FA: fractional anisotropy; ICVF: neurite density; ISOVF: isotropic volume fraction; MD: mean diffusivity; OD: orientation dispersion.

No significant genetic correlations were observed for the other neuropsychiatric conditions. This negative association with ADHD, anxiety and MDD was also apparent when using polygenic scores (PGS) in the UK Biobank (N = 53,709) (*24* ). Polygenic effects of ADHD on cortical size measures were also significant in the childhood ABCD cohort (N = 5,119), and, for global cortical SA and volume, in neonates from the developing human connectome project (dHCP, N = 455) (**Figures 1b-d, S1, Table S3**). This indicates that these shared negative genetic effects for ADHD emerge early in life, potentially prenatally. The association for ADHD is consistent with within-family epidemiological study demonstrating that reduced head circumference within the normocephalic range at birth is associated with increased odds of ADHD (*26* ).

### ADHD and a shared cortical size latent trait underlies the observed associations

There is moderate to high genetic correlation among the various cortical size phenotypes, and among the three psychiatric conditions genetically correlated with cortical size (ADHD, anxiety, MDD). Consequently, we used genomic structural equation modelling (SEM) (*20* ) to model the shared genetics among these phenotypes to assess whether a small number of latent traits underlie the observed genetic correlations.

First, we recalculated the genetic correlations between the regional cortical size phenotypes and the three psychiatric conditions after conditioning on the genetic effects of the corresponding global phenotypes, and additionally for regional subcortical volumes after conditioning on total subcortical volume. No regional cortical size phenotypes or subcortical volumes were significant at the Bonferroni-corrected significance threshold of *p <* 8.12 *×* 10^−6^, although several were nominally significant with modest effect sizes (**Figure S2, Table S4**). This indicates that global brain size accounts for most observed genetic correlations, with some regional IDPs potentially contributing weaker, independent effects.

Subsequently, we modelled the shared genetics of the four global cortical size IDPs (grey matter volume, surface area, intrinsic curvature, and folding index), given their high genetic correlation. A single common factor emerged with high loadings across all measures and good model fit, replicating our previous finding (*27* ) that these global factors reflect a single underlying cortical size factor (**Figure 2a-b, Table S5**). This factor was negatively correlated with ADHD, anxiety and depression (*r*_*g*_ = −0.21 *±* 0.04, −0.17 *±* 0.04, −0.13 *±* 0.03, respectively, **Table S6**).

**Figure 2:**
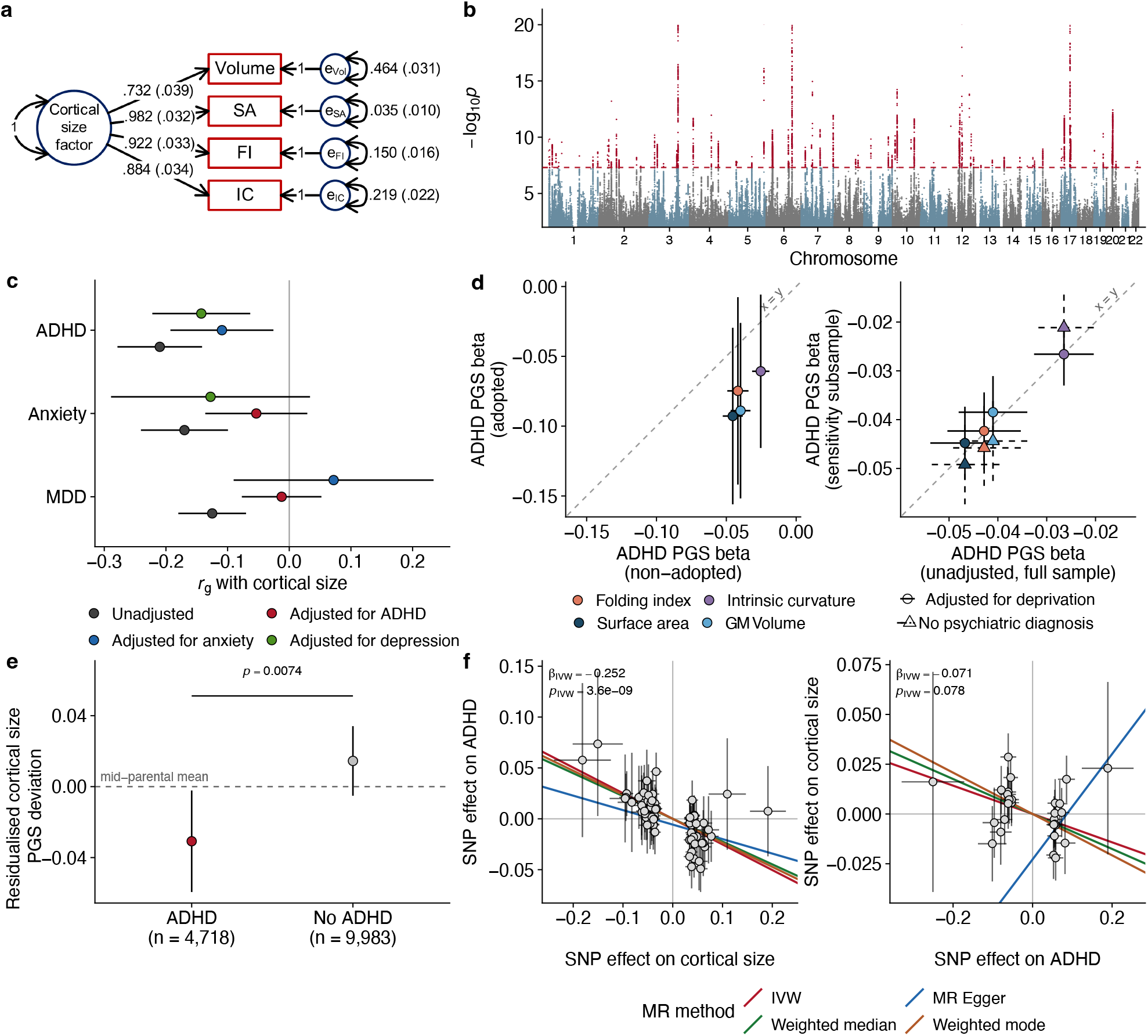
The cortical size common genetic factor. **a** Definition of the cortical size factor by genomic structural equation modelling. **b** Manhattan plot of variant-level associations with the cortical size factor. **c** Genetic correlation between cortical size and psychiatric disorders adjusted for one another. **d** Effect size estimates and standard errors from polygenic scores of ADHD to cortical size IDPs compared across non-adopted and adopted individuals (left), and comparing the base-model effect size with effect sizes either in healthy individuals only (triangles) or after adjusting for deprivation (circles) (right). Both analyses were conducted in the UK Biobank. The dashed line is *x* = *y*. **e** Deviation of cortical size polygenic score from the parental mean for ADHD and unaffected children in the SPARK dataset. **f** Mendelian randomisation scatterplots from cortical size to ADHD (left) and from ADHD to cortical size (right). Dots and crosses represent effect size and standard error of genetic variants on both phenotypes.

Finally, as there are moderate positive genetic correlations among the three psychiatric conditions, we next tested whether the genetic correlation between the cortical size factor and neuropsychiatric conditions was driven by a single condition. Using GWAS by subtraction (*28* ), we found that genetic correlations with the cortical size factor were attributable to ADHD: ADHD-independent genetic components of anxiety and major depressive disorder showed no significant genetic correlation with the cortical size factor, whereas the anxiety- and MDD-independent genetic component of ADHD remained significantly negatively correlated (*r*_*g*_ =-0.11 to -0.14, **Figure 2c, Table S6**). Together, these analyses indicate that shared genetics between a cortical size factor and ADHD explains the majority of the observed genetic correlations, particularly the shared genetics with anxiety and MDD. Hence, we focus on the cortical size factor and ADHD in subsequent analyses.

### Reduced cortical size is causally related to increased risk for ADHD

We conducted four additional analyses to contextualise the negative associations between ADHD and the cortical size factor. First, we compared the co-localisation of the effects of the ADHD PGS on surface area against established case-control maps of surface area from the ENIGMA consortium (*29*). We identified moderate and significant co-localisation between the ADHD PGS effects on the cortex and difference in SA in individuals with ADHD compared to controls (**Figure S3a**). This suggests that the effects of ADHD PGS on surface area are topographically similar to observed case-control differences in ADHD. Second, although differences in SA in ADHD has only been observed in children and not in adolescents and adults (*3*), we observed consistent negative genetic correlations using age-at-diagnosis-stratified GWAS of ADHD, suggesting that the observed genetic associations are robust to phenotypic definitions of ADHD (**Figure S3b-d, Table S7**) (*30, 31*). Third, in the UK Biobank, effects of ADHD PGS on global measures of cortical size were similar when restricted to individuals without any known psychiatric conditions, suggesting that the effects are not driven by a subset of individuals with a psychiatric diagnosis (**Figure 2d, Table S8a**). Fourth, in the UK Biobank, the effects of ADHD PGS did not differ between those adopted and not adopted; and did not attenuate after controlling for regional socioeconomic deprivation, (**Figure 2d, Table S8a**), indicating that the observed results do not entirely reflect environmental confounding. Consistent with this, in trio-based analyses, individuals with ADHD had an undertransmission of PGS for cortical size from the mid-parental mean, whilst individuals without ADHD did not (**Figure 2e, Table S8b**). This suggests that the observed effects of ADHD PGS reflect direct genetic effects.

Given the robustness of the association between ADHD and cortical size, to determine whether there is a causal relationship between the cortical size factor and psychiatric disorders, we conducted two-sample Mendelian Randomisation analyses (**Methods**). Inverse-variance weighted Mendelian Randomisation demonstrated that decreasing cortical size was causally associated with increased likelihood of ADHD. These findings were consistent when using methods that address bias due to pleiotropy (MR-Egger, MR-Presso, mode- and median-weighted MR)(**Figure 2f, Table S9a**). Furthermore, these results were also significant when using instruments derived from the global GWAS of SA, FI, IC, and GMV (**Table S9b**). We observed a less consistent causal relationship between the cortical size factor and MDD and anxiety disorder, and this relationship was no longer statistically significant after conditioning on the genetic effects of ADHD using GWAS by subtraction (**Table S9c**). We observed no evidence for causality in the opposite direction despite sufficient statistical power (F-statistic for each genetic instrument *>* 10). Overall, these results suggest that reduced cortical size is causally upstream of ADHD, anxiety and MDD.

### Cortical size and ADHD are enriched in different cell types in the developing brain

We reasoned that the association between cortical size and ADHD reflects shared cellular processes in cortical development. Genetic influences on cortical size are enriched in the proliferative capacity of neural progenitor cells, including radial glia and intermediate progenitor cells (IPCs) (*27* ). Given this, and the observation of negative association between ADHD PGS and global cortical size phenotypes in neonates from the dHCP cohort, we hypothesised that the shared genetic architecture between ADHD and cortical size will converge on either specific cell types or gene programmes active during prenatal development.

To test this, we calculated the specificity of gene expression in each cell type using adult (*32* ) and developmental (*33* ) single nucleus RNA-sequencing (snRNA-seq) data (**Methods**). Enrichment was then estimated by regressing the specificity scores for each cell type with GWAS effect sizes using MAGMA (*34* ). A significant positive enrichment indicates that genes with specific and stable expression in a cell type have lower p-values in the GWAS of ADHD or cortical size. Because MAGMA is not sensitive to the direction of effect of a gene on a trait, genes for ADHD and cortical size can be enriched in the same cell type even if they are negatively correlated: genes that increase or decrease the trait can have similarly low p-values in MAGMA gene-based analyses.

In the developing brain, cortical size-related genes were enriched for progenitor cells, whilst ADHD-related genes were enriched in differentiated excitatory neurons, particularly deep layer excitatory neurons (**Figure 3a, Table S10a**). No cell type was enriched for both traits on either the developing or adult brain (**Figure 3b, Table S10b**). In sensitivity analyses, differentiated neurons were also enriched significantly for the MDD GWAS, and nominally for the anxiety GWAS in both the developing and the adult brain. These enrichments attenuated after adjusting for the genetic effects of ADHD (**Figure S4**). These patterns were replicated using single-cell Disease Relevance Scores (scDRS, **Figure S5, Table S11**).

**Figure 3:**
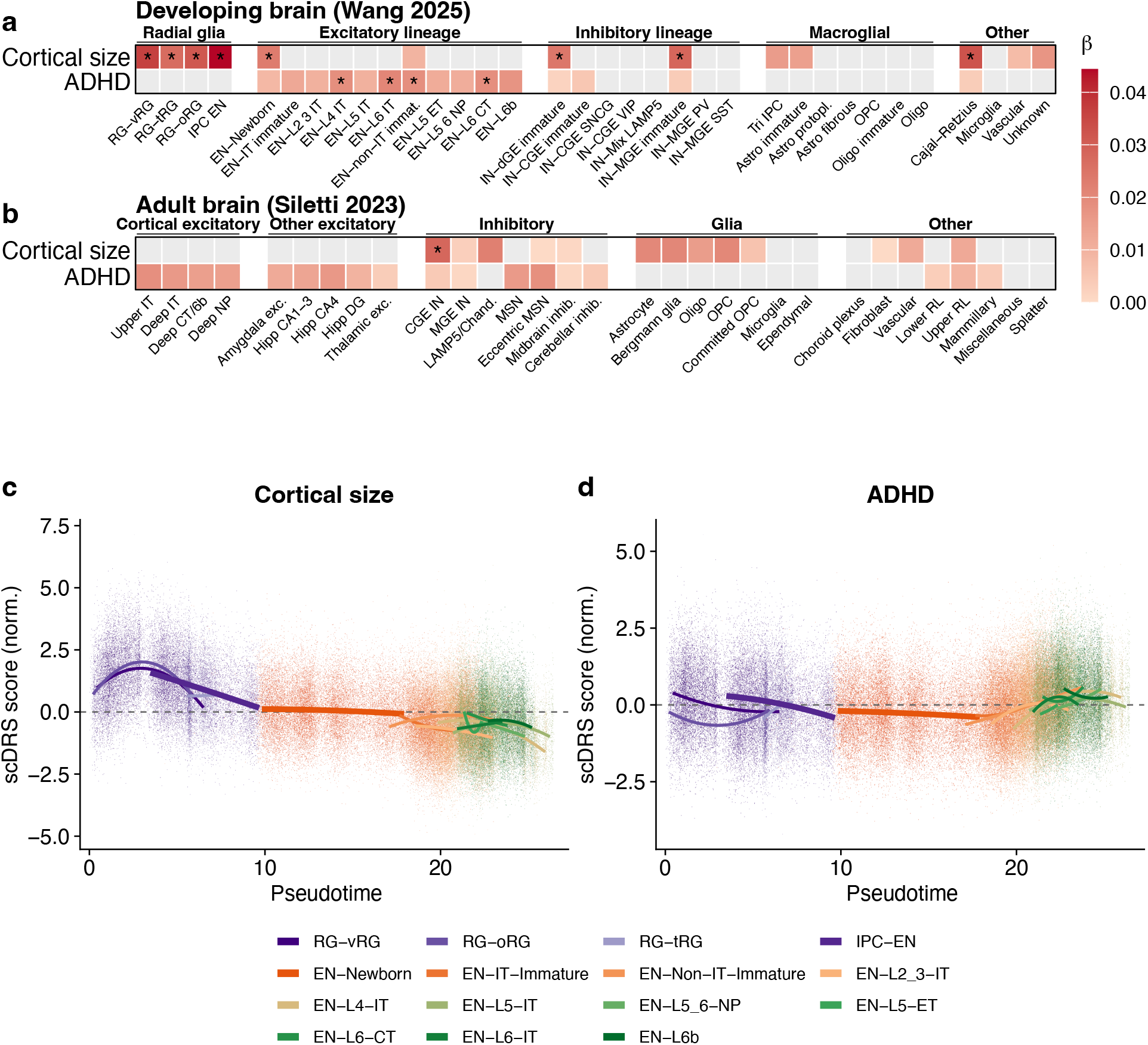
Cell-type enrichment of ADHD and cortical size. **a-b** Cell-type-level enrichment of cortical size and ADHD in (**a**) the developing brain and (**b**) the adult brain, as estimated by scoring gene expression specificity in each cell type using Cepo, and correlating with GWAS effect sizes using MAGMA. *: Passing the Benjamini-Hochberg *q <* 0.05 threshold. Non-significant (*p >* 0.05) associations are coloured grey. **c-d** Cell-level enrichment scores for (**c**) cortical size and (**d**) ADHD estimated by scDRS, plotted against pseudotime estimated for the excitatory lineage, with quadratic best-fit lines for each cell type. Colour codes of each cell type are shown below the figure. RG: radial glia; IPC: intermediate progenitor cell; EN: excitatory neuron; L2-L6: cortical layers 2-6; IT/ET: intra-/extra-telencephalic; IN: inhibitory neuron.

These findings suggest that the genetic effects of cortical size and ADHD are both enriched in foetal brain development, but in different cell populations at different stages along the progenitor-excitatory neuron lineage (**Figure 3c-d**). Consequently, we hypothesised that the polygenic effects of ADHD and cortical size will overlap in developmental genetic programmes, particularly those that regulate the differentiation of progenitors to neurons.

### Shared neurogenesis programme in mid-gestation

Biological processes typically emerge from coordinated regulation and expression of groups of genes, within and across cell types, which are referred to as gene expression programmes. Some of these processes (e.g., neurogenesis and mitosis) emerge from transient, coordinated changes in gene expression. To identify gene expression programmes, we decomposed snRNA-seq data using consensus non-negative matrix factorisation (cNMF) (*35* ). This method also allowed us to quantify the transcriptional activity of each programme in each cell. We first analysed a developmental dataset spanning the first trimester to adolescence by Wang et al (2025) (*33* ).

We decomposed this dataset into 16 gene co-expression programmes (**Methods, Supplementary Note**).

Using an ensemble classifier (**Methods**), six of these programmes were enriched for single cell types, nine were enriched across multiple cell types, and the remaining one was not enriched in any specific cell type (**Table S12**). Only one of these programmes (Wang F3) was enriched for both cortical size and ADHD GWAS (**Figure 4a, Table S13**). This factor was highly expressed in both intermediate progenitor cells (IPCs) and newborn excitatory neurons, and peaked at the transition between the two cell types along the radial glia-excitatory neuron pseudotime axis (**Figure 4b-d**). Along chronological time, the Wang F3 activity peaks at approximately post-conception week (PCW) 15 (**Figure 4e**). The top genes of Wang F3 include transcription factors involved in neuronal differentiation and corticogenesis (*EOMES, SOX11, SOX5, NFIA, NFIB, PBX1, MEIS2* ), and are enriched in GO terms such as axon guidance, *Eph*-ephrin, semaphorin-plexin and netrin signalling pathways (**Figure 4f, Table S14**). Using an independent study on IPC subtypes (*36* ), we confirmed that Wang F3 is enriched for genes up-regulated in late, neuronal-like IPCs compared to early, radial-glia-like IPCs (fold enrichment 3.92, *p <* 1 *×* 10^−5^, **Table S15, Methods**).

**Figure 4:**
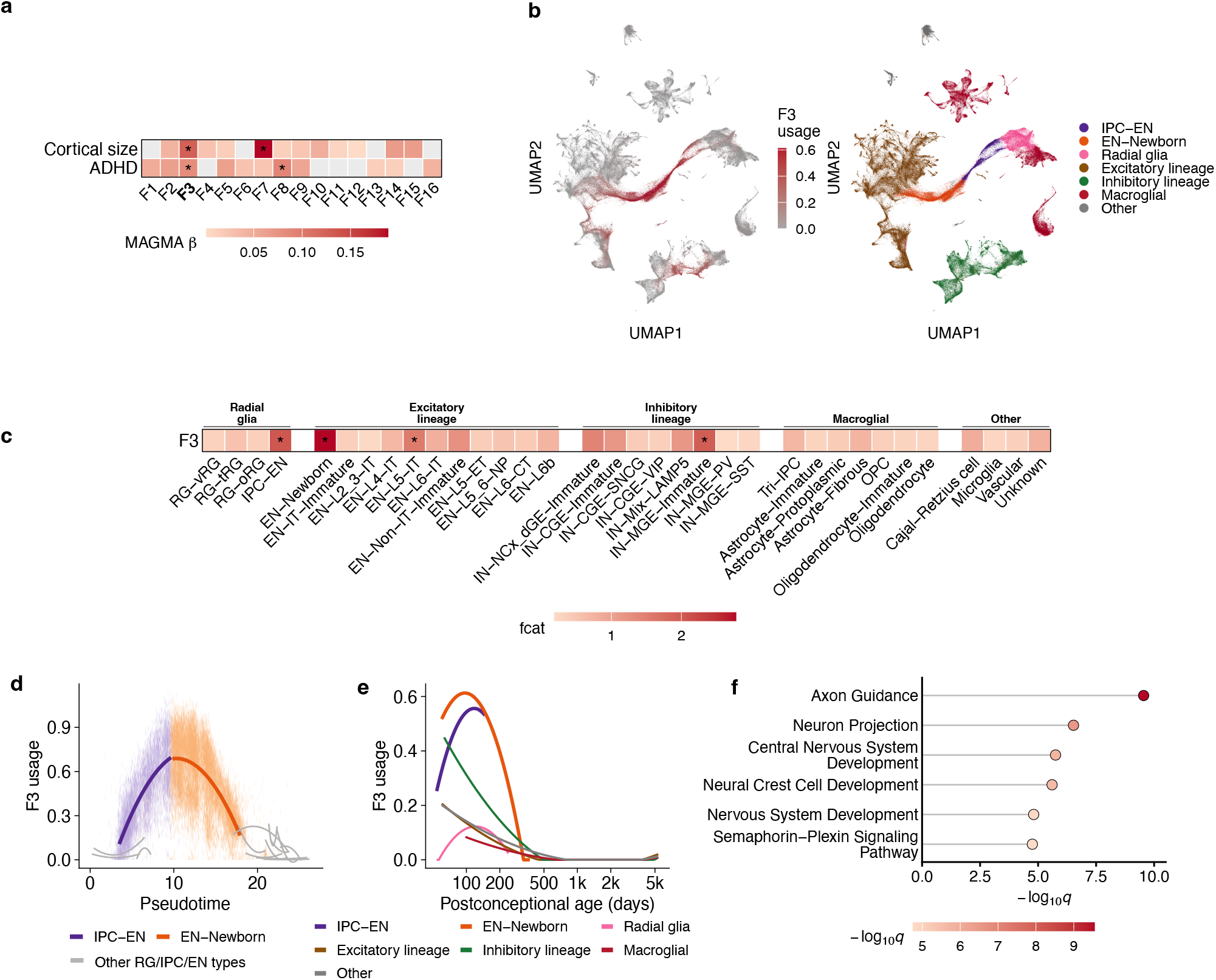
Consensus non-negative matrix factorisation (cNMF) identified gene co-expression programmes from the Wang (2025) dataset (*33* ). **a** Association between ADHD, cortical size and 16 cNMF factors using MAGMA gene-covariate analysis. *: Passing the Benjamini-Hochberg *q <* 0.05 threshold. Non-significant (*p >* 0.05) associations are coloured grey. **b** Expression activity of F3 in UMAP space. Inset: cell-type legend re-drawn using data from Wang (2025) (*33* ). **c** Cell-type enrichment score (*F*_*cat*_) of F3 as estimated using an ensemble classifier (*37* ). The significance threshold was automatically determined by the matrix of all cell-type enrichment scores across 16 gene expression factors. **d-e** Expression activity of F3 in the excitatory lineage against (**d**) pseudotime and (**e**) post-conception age, with quadratic best fit lines for each cell type. **f** Gene ontology term enrichment of the top 200 genes in F3.

To test replicability, we applied cNMF to an independent snRNA-seq dataset (PCW15-16, near the peak of Wang F3 activity). This identified a similar axonogenesis-enriched programme (Polioudakis F3), correlated with Wang F3 (r = 0.647), that replicated the overlapping enrichment for both ADHD and cortical size GWASs (**Figure S6, Table S15-17, Supplementary Note**).

### Shared progenitor programmes in the first trimester

The enrichment of deep layer neurons in ADHD (**Figure 3a**), which are primarily generated in the first trimester, suggested a role for gene programmes in the first trimester. We reasoned that the relatively low sampling from the first trimester in Wang et al (2025) (*33* ) offers limited resolution for the identification of programmes active in the first trimester. Consequently, we used the largest snRNA-seq data from the first trimester with samples from both cortical and subcortical regions by Braun et al (2023) (*36* ) to identify cNMF factors.

In the Braun dataset, we identified 18 cNMF factors (**Methods, Supplementary Note**). Four of these (Braun F1; Braun F3, Braun F6, and Braun F7) were enriched for both cortical size and ADHD (**Figure 5a, Table S18**). Braun F1 contains multiple ribosomal proteins and represents a factor related to regulation of cytoplasmic translation (**Figure S7a**). Because this factor regulates a transient activity common to almost all cell types and brain regions, we did not consider this factor further in our analyses. Braun F3 is active in radial glia only and shows broad activity spanning the midbrain, thalamus, and telencephalon. Braun F7 is active in IPCs and neuroblasts (a post-mitotic, migratory stage between IPCs and newborn neurons) and in the developing cortex and midbrain. Braun F6 is less cell type specific, with the highest activity in IPCs, and is active primarily in the subcortex and striatum (**Figures 5b, S7b-d, Table S19**). These three factors are enriched for the broad GO term “nervous system development” (**Figure 5c, Table S20**), but they represent different developmental stages in the neurogenesis process.

**Figure 5:**
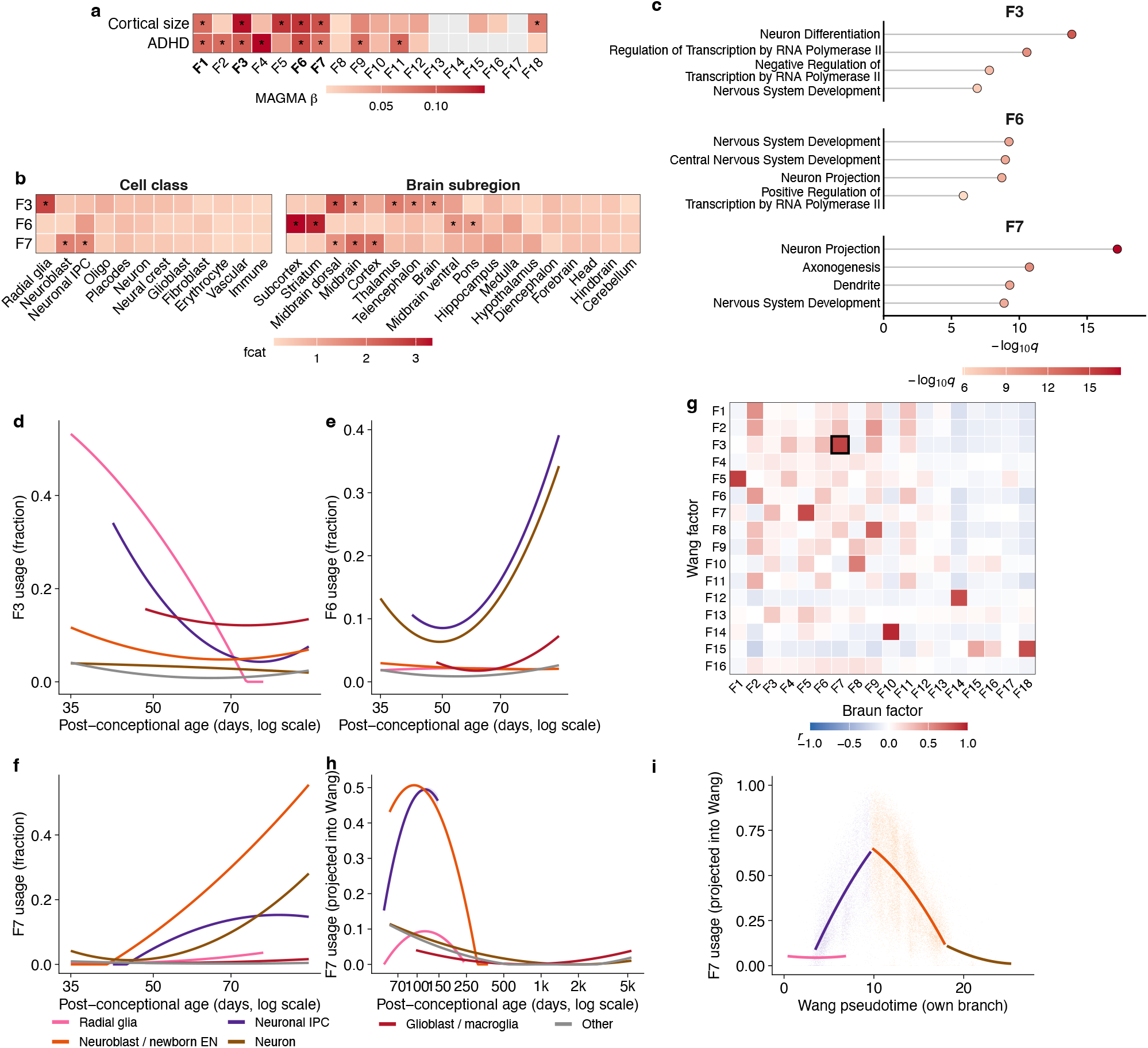
Consensus non-negative matrix factorisation (cNMF) identified gene co-expression programmes from the first trimester (Braun (2023) dataset) (*36* ). **a** Enrichment of ADHD and cortical size in cNMF factors using MAGMA gene-covariate regression. *: Passing the Benjamini-Hochberg *q <* 0.05 threshold. **b** Enrichment score (*F*_*cat*_) of F3, F6 and F7 in cell types and brain regions as estimated using an ensemble classifier (*37* ). **c** Gene ontology enrichment of the top 200 genes in F3, F6 and F7. **d-f** Expression activity of F3 (**d**), F6 (**e**) and F7 (**f** ) plotted against post-conception age, with quadratic best-fit lines for highest-expression cell types. **g** Correlation between gene-level weights of Braun and Wang factors. The highest correlated factors (Wang F3 and Braun F7) are highlighted. **h-i** Expression activity of Braun F7 projected onto the Wang dataset against (**h**) post-conception age and (**i**) pseudotime, with quadratic best-fit lines for each cell type.

The expression of Braun F3 peaks in radial glia at the earliest sampled timepoint, PCW 5 (**Figure 5d**), and was enriched for *WNT, Notch*, and *SHH* signalling pathways, with the highest-loading genes including progenitor self-renewal regulators (*CTNNB1, HES4/5, OTX2* ). Furthermore, Braun F3 is enriched for genes up-regulated in radial-glia-like IPCs (fold enrichment 2.44, *p <* 1 *×* 10^−5^, **Table S15**) but not neuronal-like IPCs, reflecting upstream competence specification. Both Braun F7 and Braun F6 are neuronal differentiation and axon guidance programmes (**Figure 5e-f, Table S20**). Expression of both programmes increases over PCW10-14, until the end of the sampled period. Braun F6 is a subcortical/striatal/thalamic programme and includes TFs associated with inhibitory fate specification (*DLX1/2/5, ARX, GSX1, GAD1/2* ). It is enriched in neuronal-like IPCs (fold enrichment 2.143, *p* = 0.003, **Table S15**). Braun F7 was mostly cortical, with additional enrichment in the midbrain, and included TFs in excitatory fate specification (*EOMES, SOX11, BCL11B, NF1A, NFIB* ) and axon guidance receptors (*ROBO2, SEMA3A/3C, EPHA3/5, DCC* ). Braun F7 factor was highly correlated with Wang F3 (Pearson r = 0.865, **Figure 5g, Table S16**), and had similar enrichments for GO terms and for genes upregulated in neuronal-like IPCs (fold enrichment 5.802, *p <* 1 *×* 10^−5^, **Table S15**). Indeed, projecting Braun F7 onto the Wang dataset identified a peak at approximately PCW14 (**Figure 5h-i, Supplementary Note**), suggesting that Braun F7 was capturing the same biological processes as Wang F3 in the first trimester.

Together, across all three snRNA-seq datasets tested, these findings suggest that the shared genetic architecture of cortical size and ADHD spans the full cascade of first-trimester to mid-gestation neurogenic commitment. Despite better statistical power, the GWAS of anxiety was only enriched for Braun F3, and the GWAS of MDD for Wang F3, Braun F7, and Braun F6 (**Figure S8, Tables S13**,**S18**). Adjusting for ADHD attenuated all enrichments, with the exception of the MDD enrichment in Braun F6. This suggests a relative specificity of these enrichments to ADHD.

Given that the top loading genes in both Wang F3 and Braun F7 contained several transcription factors (TFs) involved in neurogenesis and fate specification, we reasoned that the polygenic signals of ADHD and cortical size may be enriched for their computationally predicted downstream targets (eRegulons) (*33* ). We tested single-phenotype enrichment in eRegulons using MAGMA (*34* ) and enrichment of genetic overlap in eRegulons using GNOVA (*38* ). Out of the 66 TF eRegulons that are a top-200-loading gene of at least one of the four convergent cNMF programmes (Wang F3, Braun F3, Braun F6, Braun F7), 28 were significantly enriched (Benjamini-Hochberg *q <* 0.05) for cortical size and 30 for ADHD, including 18 for both phenotypes (**Figure S9a-b, Table S21**). These included canonical regulators of excitatory neuronal fate determination (*MEF2C, NFIA, NFIB, SOX5/11, PAX6* ). More generally, across all TFs included in the cNMF programmes, the negative genetic covariance between ADHD and cortical size within the eRegulon of a TF was correlated with its weight on Wang F3 and Braun F7 (**Table S22, Figure S9c**). In other words, the more central a TF was to these two neurogenic programmes, the more strongly its target genes showed opposing genetic effects on ADHD and cortical size. This suggests that opposing genetic effects on ADHD and cortical size are enriched in progenitor-to-neuron transition programmes. To further test this, we calculated scDRS scores for cortical size and ADHD in dividing progenitor cells, comparing those actively amplifying the progenitor pool against those committed to differentiation to neurons (**Figure S9d, Table S23**, see **Methods**). Cortical size scDRS scores were consistently higher in amplifying progenitors, whilst ADHD scores were higher in differentiating progenitors, which held across radial glia and IPC subclasses. The overlapping cNMF programmes between ADHD and cortical size also showed differential activity across neural progenitors: Braun F3 was more active in amplifying progenitors, whereas Wang F3 and Braun F7 were more active in differentiating progenitors (**Figure S9e, Table S23**). Together, this suggests that cortical size and ADHD overlap on genetic programmes regulating progenitor pool expansion versus neuronal fate commitment.

### Spatial specificity of genetic overlap

To test the progenitor-versus-neuron dissociation independently, and to establish where in the developing brain the four cNMF programmes (Wang F3; Braun F3, Braun F6, and Braun F7) and polygenic signals converge, we used gsMap (*39* ) to map the polygenic effects of cortical size and ADHD onto spatial transcriptomic sections of the mid-gestation human forebrain (N = 4 sections: one PCW12 and three PCW15, frontal cortex; **Table S24, Methods**).

Across all sections, cortical size was highly enriched in the proliferative zones, particularly in the outer subventricular zone (oSVZ), and extending into the ganglionic eminences and the intermediate zone (IZ) (**Figure 6a-c, Table S25**). Consistent with the snRNA-seq results, ADHD was enriched primarily in postmitotic and subcortical regions, but was additionally enriched in the proliferative zones at PCW12, an enrichment that was weaker by PCW15. Both traits were enriched in the oSVZ, the transient germinal zone wherein outer radial glia and other progenitors undergo the transit-amplifying divisions to expand the cortical surface area and produce neurons, and in the IZ, through which newborn neurons migrate to the cortical plate. Additionally, at PCW15, both traits overlapped in the medial ganglionic eminence. When applying the same analysis to anxiety and MDD, we did not identify consistent enrichment for either in the proliferative zones, despite better-powered GWAS. Rather, anxiety and MDD were primarily enriched in postmitotic regions, indicating that the proliferative-zone signal for ADHD reflects specificity rather than statistical power (**Figure S10**).

**Figure 6:**
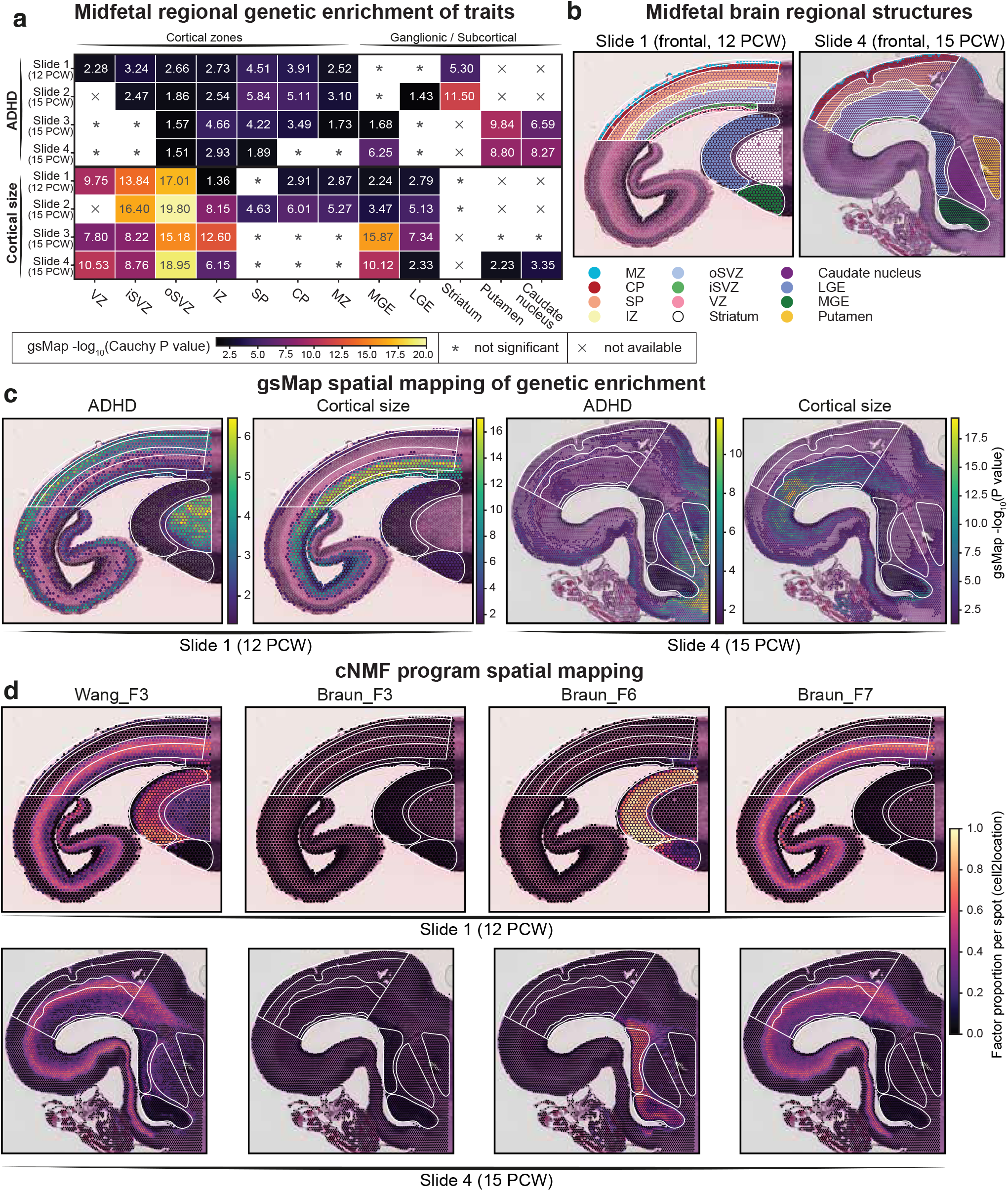
Spatial enrichment in the developing brain. **a** Enrichment of ADHD and cortical size in different brain regions, as estimated by Cauchy combination of spot-level enrichment using gsMap. **b** Brain region annotation of sections 1 and 4. Un-coloured spots are not assigned to a particular anatomical region. White lines indicate the boundary between regions. The same boundaries are drawn in panels **c-d** for reference. **c** Spot-level enrichment p-values (in log scale) of ADHD and cortical size estimated using gsMap. Non-significant (*p >* 0.05) spots are not coloured. **d** Spot-level activity of cNMF programmes (Wang F3, Braun F3, F6, F7), in terms of proportion of total spot-level gene expression, estimated using cell2location (see **Methods**). For panels **b-d**, the Nissl-stained microscopy images of sections 1 and 4 are shown in the background. VZ: ventricular zone; iSVZ/oSVZ: inner/outer subventricular zone; IZ: intermediate zone; SP: subplate; CP: cortical plate; MZ: marginal zone; MGE/LGE: medial/lateral ganglionic eminence.

We next used cell2location to spatially localise the four cNMF factors on the four midgestation human forebrain sections. Wang F3 and Braun F7 both localised to the oSVZ, extending into the IZ, with high spatial correlation (r = 0.82, pooled Visium spots, *N* = 129, 625), consistent with their transcriptional overlap (r = 0.865). Braun F3 was not detected, consistent with its peak in the first trimester (PCW5-7). Braun F6 primarily localised in the subcortical ganglionic eminences (**Figure 6d**), including the medial ganglionic eminence which is enriched for the GWAS signal of both ADHD and cortical size. This is consistent with the high loading of TFs with roles in inhibitory fate specification in this programme.

The polygenic signals for ADHD and cortical size, and the cNMF programmes enriched for the two sets of polygenic signals, therefore converge at the same anatomical location: the oSVZ and IZ in the cortex where progenitor amplification leads to postmitotic differentiation and migration, and the medial ganglionic eminence in the subcortex, which gives rise to cortical inhibitory interneurons. This spatial counterpart of the cell-transition programmes identified in snRNA-seq provides convergent evidence for the role of neurogenic programmes active in the midgestation cortex and subcortex underlying the negative genetic correlation observed between cortical size and ADHD.

### Convergence with human cortical evolution

The oSVZ has undergone disproportionate expansion in humans, and this, combined with the increased proliferative capacity of basal radial glia in the oSVZ is thought to contribute to the marked expansion of the cerebral cortex over the course of human brain evolution (*40* –*42* ). Given the overlapping enrichment of cortical size, ADHD and their underlying neurogenic genetic programmes in oSVZ (**Figure 6**), we asked whether this genetic overlap also converges with molecular signatures of human-specific cortical evolution.

We first tested whether regional genetic correlations between ADHD and cortical surface area was spatially aligned with an axis of evolutionary cortical expansion using spin-based permutation. The regional genetic correlation of the surface area of ADHD was significantly spatially correlated with regions of the brain that have expanded in humans compared to macaques (Pearson *r* = −0.491, Benjamini-Hochberg *q* = 0.0014 using spin permutation, **Figure S11a, Table S26**). In other words, cortical regions showing the greatest expansion in humans compared to macaques had the strongest negative genetic correlation with ADHD and cortical size measures. Comparison with other topographic brain maps indicated that this spatial co-localisation is most consistent with evolutionary cortical expansion. Sensitivity analyses using genetic correlation between ADHD and other measures of cortical size (volume, folding index, intrinsic curvature) showed directionally consistent results (**Figure S11a, Table S26**).

We next asked whether the genetic overlap between ADHD and cortical size was enriched for genes differentially expressed (DEGs) in humans compared to macaques using snRNA-seq data from the developing cortices, and additionally DEGS identified from human and chimpanzee cortical organoids (*43* ). Using MAGMA, we found that both GWASs were enriched for human-macaque DEGs shared across cell types, as well as human-macaque DEGs specific to IPCs and inhibitory neurons, but not human-chimpanzee DEGs (**Figure S11b, Table S27**). GWAS for MDD but not anxiety was also enriched in the human-macaque DEG sets, but this attenuated and was no longer statistically significant after conditioning on the genetic effects of ADHD. Focusing on human-macaque DEGs for subsequent analyses, GNOVA (*38* ) showed that ADHD and cortical size share significant negative genetic covariance within DEGs shared across all cell types (*ρ* = −0.0027, *p* = 0.0142), and within IPC-specific DEGs (*ρ* = −0.0032, *p* = 0.0025, **Table S28**).

We next tested whether the four convergent cNMF programmes were also enriched for the human-macaque DEGs with a permutation t-test on the cNMF weights. All four shared programmes (Wang F3, Braun F3, Braun F6, Braun F7) showed significant enrichment (**Figure S11c, Table S29**). Because DEGs may be highly loading across multiple factors, we tested whether these DEGs were more strongly enriched in the four factors compared to other cNMF factors, by normalising the cNMF weights across all factors. Whilst the radial glia factor (Braun F3) was no longer enriched in DEGs in any cell type, Wang F3 and Braun F7 were enriched for DEGs shared across cell types as well as DEGs specific to IPCs and excitatory neurons, consistent with their activity in excitatory neurogenesis and localisation to oSVZ and IZ. Braun F6 was enriched in DEGs across all cell types, and DEGs specific to inhibitory neurons (**Figure S11d, Table S29**), consistent with its activity in inhibitory fate specification and localisation to ganglionic eminences.

Together, these findings indicate convergence between the genetic architecture of ADHD and cortical size and signatures of human cortical evolution at both anatomical and molecular levels. The overlap suggests that the developmental programmes linking cortical size and ADHD intersect with biological mechanisms that differ between humans and macaques.

## Discussion

Structural brain differences have been described in neuropsychiatric conditions for several decades, but elucidating the mechanisms that underlie these has proven challenging. Here, by combining neuroimaging with population and functional genetics, we demonstrate a causal link between reduced cortical size and ADHD that converges on a mid-gestation neurogenic programme. Cortical size and ADHD are enriched in different cell types, proliferating progenitors and postmitotic excitatory neurons, respectively, with no cell type enriched for both. Nevertheless, they converge on a gene programme active at the developmental transition between these cell types, which peaks in mid-gestation and is localised to the proliferative outer subventricular zone and the transitory intermediate zone. These findings, replicated across three independent datasets and using both single-nucleus and spatial transcriptomic data demonstrate that polygenic effects can converge on molecular mechanisms not captured by canonical analyses that map genetic risk to cell types. This programme is also enriched for genes differentially expressed between human and macaque cortex, particularly among those specific to intermediate progenitors and excitatory neurons.

The observation that ADHD is associated with smaller cortical size and related measures within the normocephalic range is not new, and has been reported extensively by epidemiological and MRI studies (*3, 44* ). To contextualise the scale of this association here, a one-SD increase in ADHD PGS was associated with a *∼*0.05SD reduction in global cortical surface area in adults, children, and neonates - a small but consistent effect from before birth into adulthood. Here we show that this is partly due to shared genetics, and this genetic association is already present in neonates before the onset of ADHD symptoms. Under-inheritance of cortical size polygenic scores in individuals with ADHD, alongside equivalent effects of ADHD polygenic scores in adopted and non-adopted individuals, and before and after conditioning on socioeconomic status, all together indicate that this relationship cannot entirely be attributed to familial and environmental confounding, but instead captures direct genetic effects. Additionally, Mendelian randomisation supports a causal effect of reduced cortical size on ADHD, but not *vice versa*, despite adequate statistical power.

At the molecular level, although we are unable to directly test causality, three observations suggest that there are opposing polygenic effects of ADHD and cortical size along the neurogenic transition. First, among dividing progenitors, those with higher expression of ADHD-related genes were more likely to be committed to neuronal differentiation than to be amplifying the progenitor pool, with opposing effects for cells with higher expression of cortical-size related genes; this held across progenitor subclasses. Second, the polygenic effects of the downstream targets of transcription factors in the overlapping programmes are opposing for ADHD and cortical size, and the strength of this opposition correlates with how strongly the transcription factor loads onto the programmes. Third, the convergent programmes themselves lie along this axis: Braun F3 is a radial glial self-renewal programme, with high loadings for regulators of progenitor proliferation (e.g., *CCND1, HES4/5, CTNNB1, OTX2* ) but not neuronal or laminar identity genes. In contrast, Wang F3, Braun F7 and Braun F6 are neurogenic fate-commitment programmes with high loadings for intermediate progenitor and newborn neuron markers (*INSM1, SOX4/11, DCX, MYT1L*), carrying lineage-specific fate determinants for excitatory (*EOMES, NEUROD2/6, PPP1R17* ) and inhibitory (*DLX1/2/5, GAD1/2, ARX, LHX6* ) identity, and acting in the excitatory (Wang F3, Braun F7) and inhibitory (Braun F6) lineages respectively. Together, these suggest a developmental switch: polygenic effects for cortical size bias progenitors towards proliferation, increasing the progenitor pool and thereby cortical size, whilst polygenic effects for ADHD bias progenitors towards neuronal fate commitment. These observational findings have preliminary, albeit mixed, support from model systems. Neural stem cells derived from individuals with ADHD proliferate more slowly than controls, an effect absent at the pluripotent stage (*45* ), and telencephalic organoids derived from an individual with ADHD show altered early cortical layer structure (*46* ). Another hiPSC study did not observe differences in progenitor proliferation but reported dysregulated glutamatergic neuronal development in ADHD compared to controls (*47* ). Definitive testing will require larger-scale iPSC panels differentiated to cortical progenitors, in which the effects of PGS of ADHD and cortical size on the progenitor-to-neuron transition can be observed.

The convergent programmes differ in developmental timing and lineage. Braun F3 peaks in the first trimester, coinciding with deep-layer neurogenesis. It was undetectable in our PCW12 and PCW15 sections, so its spatial localisation is unknown. Wang F3 and Braun F7, which are effectively the same programme in two different datasets, peak at PCW12-15 in the outer subventricular and intermediate zones, with high loadings from intratelencephalic and upper-layer identity markers (*CUX2, UNC5D, SATB2, BCL11A, PTPRK, LHX2* ). Deep-layer markers (*BCL11B, SOX5* ) are also present but weakly loaded. Braun F6 shows the same transition architecture in the inhibitory lineage (*DLX1/2/5/6, GAD1/2, ARX, LHX6, SP9* ), restricted to the ganglionic eminences.

This laminar character of the Wang F3/Braun F7 and Braun F6 programmes contrasts with the deep-layer enrichment of ADHD genetic signal found in snRNA-seq data, and suggests at least two temporally distinct mechanisms. An early, progenitor-competence phase captured by Braun F3 coincides with the emergence of deep-layer corticofugal neurons in the first trimester. This likely contributes to the development of the corticostriato-thalamo-cortical circuits that have been previously implicated in ADHD (*48* ) and are targeted by stimulant medications (*49* ). A second mid-gestation phase captured by Wang F3/Braun F7, localises at the oSVZ and in the IZ and is involved in fate restriction. These programmes are enriched for upper-layer identity markers and may possibly contribute to the development of intratelencephalic neurons and additional corticostriatal projections. In parallel, Braun F6, which peaks in midgestation, contributes to fate restriction of inhibitory interneurons. Consistent with this, ADHD enrichment extends to the ganglionic eminences and basal ganglia.

The overlap between the top genes in these programmes, particularly Wang F3 and Braun F7, with genes differentially expressed in human and macaque developing cortices provides correlative evidence that molecular and cellular mechanisms that have evolved along the human lineage may be associated with ADHD. Wang F3 and Braun F7 are active in the oSVZ, which primarily gives rise to upper-layer neurons, whose expansion is thought to account for much of the increase in human cortical surface area (*40* ). It is possible that several of these genes are involved in neurogenic cell fate, and also alter the neurogenic tempo. Compared to other primates, human brain development, including neurogenesis occurs at a slower tempo (*50, 51* ). ADHD-derived telencephalic organoids show a shift towards neurogenic divisions and reduced proliferation compared to control organoids, consistent with accelerated neurogenic tempo (*46* ). Pertinent to this, a recent study on cortical slice culture models of Down syndrome, which is associated with both increased odds of ADHD and smaller brains (*52, 53* ), demonstrated accelerated neurogenic tempo (*54* ). Several of the genes that overlap between human-macaque DEGs and Wang F3/Braun F7 are regulators of progenitor differentiation (e.g., SOX4 and SOX11), consistent with a potential role in neurogenic tempo. Individuals with ADHD often face challenges in complex executive function, attention and processing speed, aspects of which are thought to be uniquely developed in humans (*55, 56* ), and absolute brain volume correlates with impulse-control performance across animals, including within non-human primates (*57* ). Whether the same cortical developmental programmes that alter the neurogenic tempo and cell fate in mid-gestation, also shape these cognitive capacities, contributing to ADHD remains unknown.

More broadly, these findings provide one framework for identifying biological mechanisms underlying psychiatric conditions. Mechanistic understanding of ADHD has been shaped largely by pharmacology: stimulant medications act on catecholaminergic signalling, and this has anchored aetiological models for decades (*49* ). However, the mechanism through which common genetic variation confers risk has remained largely unresolved (*58* ). By leveraging shared genetics with an endophenotype (cortical size) whose developmental biology is comparatively well established (enriched for progenitor proliferation) (*27* ), we identify a mechanism through which some of the polygenic effects act across cell types, gene programmes and development.

The effects we describe are modest. Genetic correlations and covariances between cortical size and ADHD are small, and this mechanism can account for only a fraction of the ADHD liability. It is therefore only one contributing cause to ADHD, consistent with the heterogeneity in this condition. It is unclear whether individuals whose liability for ADHD is driven by different molecular mechanisms differ in their presentation and outcomes. These observations are largely specific to ADHD. Although we observed similar negative genetic correlations between anxiety and MDD and cortical size phenotypes in adults, these are no longer significant when conditioning on the genetic effects of ADHD. This attenuation is also observed with Mendelian randomisation, and in enrichment analyses in single-nucleus and spatial transcriptomic data. Furthermore, polygenic scores for anxiety and MDD are not associated with cortical size differences in neonates. More broadly, the absence of genetic effects underlying well-documented MRI findings is puzzling. For example, larger ventricles and reduced hippocampal volumes in schizophrenia have been widely reported (*5* ), yet we do not find significant genetic associations despite reasonable statistical power for the corresponding GWAS. There are several potential explanations for this and other null findings, including heterogeneity within psychiatric conditions, effects driven by low-frequency and rare genetic variants, and differential polygenic effects across the phenotypic distribution. These also do not preclude the possibility that observed MRI associations in psychiatric conditions are a consequence of illness or reflect non-genetic causes, neither of which has been fully controlled for in most MRI studies.

## Methods

### Dataset

The UK Biobank (UKB) is a prospective cohort study of over 500,000 participants aged 40-69 years old at recruitment. Of these individuals, 100,000 will undergo brain scanning. The ABCD study is an ongoing cohort study of childhood and adolescence. 11,880 participants from the general population were recruited from all over the United States across 21 sites, and brain MRI was acquired for the large majority of these as part of the core study protocol. In both cohorts, we focussed on participants of genetically inferred European ancestries with quality-controlled MRI and genotype data (*N*_*max*_ UKB = 53,709; *N*_*max*_ ABCD = 5,119). The developing Human Connectome Project (dHCP) is a cross-sectional cohort of neonates scanned at term-equivalent age at the Evelina London Children’s Hospital, recruited from the neonatal intensive care unit and from healthy term-born controls (*19* ). After genetic and MRI quality control, we had a maximum of 455 participants.

### Genetic quality control

The UKB team has described in detail their genetic quality control and imputation elsewhere (*16* ). Subsequently, we included only participants of self-identified European ancestry and removed individuals who were *±* 5s.d. from the means of the first two genetic principal components (PCs), resulting in a relatively genetically homogeneous population. Additionally, we excluded participants with excessive genetic heterozygosity and those who had a mismatch between reported and genetic sex. We excluded single-nucleotide polymorphism (SNP) with minor allele frequency *<* 0.001, and that were not in Hardy-Weinberg equilibrium (*p <* 1 *×* 10^−6^), and an imputation score *r*^2^ *<* 0.4, resulting in 15,916,802 SNPs.

QC of ABCD is detailed elsewhere (*59* ). Briefly, pre-imputation we removed SNPs with genotyping rate *<* 90% and deviations from Hardy-Weinberg equilibrium (*p <* 1 *×* 10^−6^). We removed individuals with genotyping rate *<* 95%, a mismatch between genetic and reported sex, and excessive heterozygosity. Because Hardy-Weinberg and heterozygosity are miscalculated in ancestrally mixed populations, these steps were conducted within genetic ancestral groups identified by principal-component clustering after merging with 1000 Genomes phase 3 data. HWE filtering and removal of individuals with excessive heterozygosity (*±*3 s.d.) were then repeated within each identified population, relatedness and PCs recalculated on the cleaned data, and the data merged, phased (Eagle v2.4) and imputed (Minimac4) via the TOPMed Imputation Server (*60* –*62* ). Imputed SNPs with *r*^2^ *<* 0.4 or minor allele frequency *<* 0.001 were removed. For the present analyses we restricted to individuals of non-Finnish European ancestry (identified by clustering), additionally excluding those *±*5 s.d. from the mean of the first two genetic PCs within this group, giving *N*_max_ ABCD = 5,119.

For dHCP, genotypes were called from the Illumina Omni5-4 array and processed with a dedicated pipeline (https://github.com/yh464/dhcp_genotype_qc). Pre-QC mapped variant IDs to rsIDs using the official Illumina Omni5-4 v1.2 support file, ENSEMBL REST and dbSNP v157 for any IDs still missing chromosome/position after this, with indel alleles resolved by querying the longer and shorter alleles from ENSEMBL REST in place of the array’s D/I encoding; variants with IDs still missing after this were removed. Pre-imputation QC removed variants with per-variant missingness *>*0.1 or Mendelian error rate *>* 0.1, and samples with per-sample missingness *>* 0.05, Mendelian error rate *>* 0.05 per trio, or a failed sex check. Ancestry clustering was performed by merging with the 1000 Genomes reference panel (a three-pass merge: direct merge, strand-flip and re-merge of non-matching variants, then removal of any remaining problematic variants), extracting common variants (MAF*>* 0.01), estimating genetic PCs in 1000 Genomes and projecting dHCP onto them, and embedding the first 5 PCs in 2-dimensional UMAP space (fit on 1000 Genomes only, using the same transform for dHCP) to identify ancestry clusters. Within each identified ancestry, a second round of QC removed individuals with missingness *>* 0.1 or variants deviating from HWE (*p <* 1 *×* 10^−6^), and subjects and variants common to all ancestries were merged. Variants were checked against the HRC reference panel with strand-flip/allele-switch correction and formatted to VCF for imputation on the TOPMed Imputation Server (*60* –*62* ). Post-imputation, variants with *R*^2^ *<* 0.6 were removed, chromosomes concatenated and lifted back to hg19, and data separated by ancestry. We included participants of European ancestry and filtered variants by minor allele frequency *>* 0.001 within this subset (*N* = 455).

### Image acquisition

For UKB, Structural magnetic resonance images of the whole brain were acquired on a standard Siemens Skyra 3T running VD13A SP4 (as of October 2015), with a standard Siemens 32-channel RF receive head coil (*17* ).

The acquisition took place with four identical scanners at four different dedicated imaging centres (Cheadle, Reading, Newcastle, and Bristol). We utilised minimally processed T1 and T2-FLAIR MRI data from UKB application 20904. T1-weighted structural imaging followed these parameters: 1.0 mm isotropic resolution, TR = 2,000 ms, TE = 2.01 ms, TI = 880 ms and flip angle 8 degrees, and Duration = 5 minutes. T2-weighted FLAIR structural imaging followed these parameters: 1.0 *×* 1.0 *×* 1.1 mm resolution, TR = 5,000 ms, TE = 395.0 ms and TI = 1,800 ms, and Duration = 6 minutes.

For ABCD (https://github.com/nih-fmrif/abcd_protocols) (*18* ), T1-weighted imaging (1.0 mm isotropic resolution) was obtained using the following parameters: TR = 2500 ms, TE = 2.88 ms, TI = 1060 ms, flip angle 8 degrees; T2-weighted imaging (1.0 mm isotropic resolution) was obtained using the following parameters: TR = 3200 ms, TE = 565 ms, flip angle variable; and diffusion-weighted imaging (1.7 mm isotropic resolution) was obtained using the following parameters: TR = 4100 ms, TE = 88 ms, flip angle 90 degrees, 500 (6-dirs); 1,000; (15-dirs) 2,000; (15-dirs) and 3,000 (60-dirs).

For dHCP (*19* ), T2-weighted and T1-weighted structural images were acquired on a 3T Philips Achieva fitted with a dedicated neonatal 32-channel phased-array head coil, sited within the neonatal intensive care unit at the Evelina London Children’s Hospital, without sedation. T2-weighted images were acquired as multi-slice fast spin-echo sequences in sagittal and axial slice stacks (TR = 12 s, TE = 156 ms, SENSE factor 2.11 axial/2.58 sagittal), and inversion-recovery T1-weighted images with TR = 4,795 ms, TI = 1,740 ms, TE = 8.7 ms (SENSE factor 2.26 axial/2.66 sagittal), both with in-plane resolution 0.8 *×* 0.8 mm^2^ and acquired at 0.8 *×* 0.8 *×* 1.6 mm resolution before being upsampled to 0.5 mm isotropic after reconstruction. Neonates were scanned at term-equivalent postmenstrual age.

### Image pre-processing and parcellation

The T1-weighted images from UK Biobank and ABCD were further preprocessed using FreeSurfer version 6.0.1, following the standard recon-all pipeline (*63* ). This preprocessing included steps such as motion correction, non-uniformity correction, Talairach, intensity normalization, skull-stripping, and automatic tissue and subcortical segmentation. For both ABCD and UK Biobank (UKB) tabular data, we computed bilateral averages across Glasser regions (*25* ). Regional values were averaged across hemispheres, resulting in 180 cortical and 20 subcortical regions. Whole-brain values were calculated by taking the total or mean of regional values. Values greater than 5 absolute standard deviation or median absolute deviations were removed, and all IDPs were converted to z-scores for analyses. The same processing was applied to the Desikan-Killiany (DK) atlas to enable spatial co-location analyses with ENIGMA case-control maps (*29* ).

Diffusion-weighted images from UK Biobank and ABCD have been corrected for eddy currents and echo planar imaging distortion (*17, 18* ). We subsequently estimated neurite orientation dispersion and density indices (NODDI) parameters using the accelerated microstructure imaging via convex optimization (AMICO) pipeline (*64* ). Regional estimates of NODDI parameters, mean diffusivity (MD) and fractional anisotropy (FA) were obtained by co-registering diffusion-weighted with T1-weighted structural images using FSL FLIRT, and applying the AFNI 3dROIstats function across Glasser regions (*25* ). Regional phenotypes were bilaterally averaged, and global phenotypes were defined as the average across all regions.

For 180 bilaterally averaged cortical regions, we estimated 12 imaging-derived phenotypes (IDPs): volume, surface area (SA), folding index (FI), intrinsic curvature (IC), Gaussian curvature (GC), mean curvature (MC), cortical thickness (CT), mean diffusivity (MD), fractional anisotropy (FA), orientation dispersion (OD), intracellular volume fraction / neurite density (ICVF), and isotropic volume fraction (ISOVF), resulting in 12 global and 2,159 regional IDPs (one regional IDP was excluded due to low variance). We estimated the volume of 20 subcortical regions and 5 phenotypes (MD, FA, OD, ICVF, ISOVF) for 27 white matter tracts. This resulted in a total of 2,326 IDPs.

Because we only used global cortical volume and surface area from dHCP, we employed the standard dHCP structural pipeline output (*19* ) without custom pre-processing. This included motion correction, Talairach, bias correction, brain extraction, and a neonate-dedicated tissue segmentation algorithm (Draw-EM).

### Multiple testing correction

We used matrix decomposition of the phenotypic correlation matrix of all 2,326 IDPs to identify 1026 independent phenotypes (*65* ), resulting in 6,156 (1,026 IDPs *×* 6 disorders) total tests conducted for the brain-wide PGS and genetic correlation analyses. Subsequently, we used a Bonferroni corrected threshold of 8.12 *×* 10^−6^ for these analyses. For all other analyses, we used Benjamini-Hochberg based false discovery rate (FDR) adjustment for multiple testing correction.

### Genome-Wide Association Analyses and genetic correlation

We used previously generated GWAS summary statistics for all 2,326 IDPs analysed in this study from the UK Biobank (*N* = 53,709) (*24* ). All GWAS were generated using the quality controlled genetic and phenotypic dataset using linear mixed-effects models employed in fastGWA (*66, 67* ). We included the following covariates: 20 genetic principal components, scan site (for UKB only), sex, age, age^2^ age *×* sex, age^2^ sex. We obtained external genome-wide association study (GWAS) summary statistics for six well-powered neuropsychiatric conditions (*58, 68* –*72* ): attention deficit hyperactivity disorder (ADHD), autism, anxiety, bipolar disorder, major depressive disorder (MDD), and schizophrenia. Details of GWAS summary statistics are provided in **Table S1**. We conducted genetic correlation between the IDPs and the neuropsychiatric conditions using LDSC (*22, 23* ), using LD weights from the North-west European population.

### Brain-wide polygenic score analysis and sensitivity analyses

We generated polygenic scores (PGS) for the same six disorders using summary statistics that excluded individuals from the UK Biobank and ABCD in the quality controlled UKB and ABCD datasets. PGS were computed using PRScs (*73* ) with default parameters (*a* = 1, *b* = 0.5, *ϕ* = 0.01).

For all 2,326 IDPs, a mixed-effect regression is done with disease-specific PGS as the predictor and IDP as the outcome. We use the same set of covariates as the GWAS analyses above. For dHCP in particular, because participants were scanned within 2 months of age, age was defined by days post-conception, and we included the foetal age at birth as an additional covariate. Because different datasets employed different imaging protocols, we included different imaging covariates: Euler index, mean and max framewise displacement (FD) for UKB and ABCD, and an expert-derived radiology score provided by dHCP (*19* ). We then tested the spatial correspondence between regional effect estimates of polygenic risk scores and case-control difference maps from ENIGMA (*29* ) using a spherical spin permutation test (*74, 75* ).

In the UK Biobank, we performed some sensitivity analyses to assess potential environmental, socioeconomic, and disease-related confounding. First, analyses were stratified by adoption status (adopted: *N*_max_ = 798). Adoption separates genetic relatedness from the postnatal family environment; therefore, comparing PGS associations between adopted and non-adopted participants allowed us to assess the potential influence of environmental factors shared with biological parents. Second, we repeated the analyses adjusting for the Index of Multiple Deprivation (IMD) to account for area-level socioeconomic confounding. Comparing the IMD-adjusted and unadjusted estimates allowed us to test whether the observed associations were robust to socioeconomic factors that relate to both genetic liability to disorders and brain structure. Third, we restricted the analyses to healthy participants without any ICD-F diagnosis (*N*_max_ = 42, 144) and compared the resulting estimates with those obtained in the full sample, assessing whether the associations were influenced by the presence of psychiatric disorders. Together, these analyses assessed the robustness of PGS-imaging associations to familial environmental factors, socioeconomic disadvantage, and psychiatric diagnoses.

Additionally, to investigate the direct genetic effects of cortical size PGS on ADHD, we used data from 14,701 trios from the Simons Foundation Powering Autism Research for Knowledge (SPARK) cohort (iWES2 and iWES3 releases - inverse variance meta-analyses) (*76* ). Genetic QC of this cohort is described elsewhere (*77* ). This cohort has been ascertained for autism, but parent-reported information on a medical diagnosis of ADHD is available for both autistic individuals and their non-autistic siblings. We used the polygenic transmission disequilibrium test (*78* ) to generate deviation in PGS for probands from the mid-parental mean, which captures genetic effects not confounded by familial factors or population stratification, and regressed this against ADHD diagnosis after covarying for sex, age, intellectual disability, and autism diagnosis status.

### Genomic structural equation modelling (SEM)

We used Genomic SEM (*20* ) to investigate the multivariate genetic structure across imaging and psychiatric phenotypes. For the cortical size latent factor, we used a common factor GWAS model where each variant loads onto a common factor which loads onto volume, surface area, folding index and intrinsic curvature. To adjust for global IDPs when investigating the genetic correlation between regional IDPs and psychiatric disorders, we used a GWAS-by-subtraction (*28* ) design to extract the global-independent component of each regional IDP, and correlated this component with psychiatric disorders. The same GWAS-by-subtraction design was used to adjust between disorders and correlate with the cortical size factor.

### Mendelian randomisation (MR)

For imaging signatures and neuropsychiatric conditions that were significantly genetically correlated, we investigated whether there was a causal relationship between them using MR. Independent loci were identified by clumping at the genome-wide significance threshold (*p <* 5 *×* 10^−8^) using a linkage disequilibrium (LD) window of 1 Mb to identify genetic instruments for MR analysis. Significant causal relationships were identified if following criteria were met:

- MR by inverse variance weighting (MR-IVW), MR corrected for sample overlap (MR-lap) and MR by weighted median are all significant at FDR *<* 0.05 (*21, 79* ).
- If outliers were identified by MR-PRESSO, the outlier-corrected MR results were still significant at FDR *<* 0.05 (*80* ).
- MR-Egger showed effect size in consistent direction. We did not require MR-Egger to be FDR-significant because this method has lower statistical power (*21* ).
- If Steiger directionality test flagged a causal relationship for false direction, above analyses were repeated after performing Steiger filtering on the genetic instruments, and significant associations identified using the same criteria.

We conducted bi-directional MR testing the effects of cortical size (both the common factor and individual IDPs) on ADHD and vice versa in the primary analyses. In the secondary analyses, we conducted similar bidirectional MR testing the effects of cortical size on MDD, Anxiety, and both MDD and Anxiety after adjusting for the genetic effects of ADHD.

### Cell type enrichment analysis

We combined GWAS summary statistics with single-nuclear RNA sequencing (snRNA-seq) datasets to identify cell types enriched for cortical size and psychiatric disorders. For developmental brain data, we used snRNA-seq data from Wang et al (2025) (*33* ) which contained 38 samples from 27 individuals spanning PCW8 to 13 years of age, totalling 232,328 nuclei. Only prefrontal and visual cortical regions were sampled. For adult brain data, we used snRNA-seq data from Siletti et al (2023) (*32* ) which contained 105 samples from 4 individuals spanning 18 to 68 years of age, totalling 3,369,219 nuclei. Samples were taken from 10 cortical and sub-cortical regions. We used cell-type annotations in the respective datasets to estimate enrichment. We selected the following methods based on their performance in a previous benchmarking study of cell-type association methods (*81* ).

We first estimated the gene-level effect sizes for all phenotypes using MAGMA (*34* ). A 10-kb window up- and downstream was used when mapping variants to genes. On that basis, we used two parallel methods to estimate cell-type enrichment: To map snRNA-seq to gene-level effect sizes, we used Cepo to derive expression specificity scores of each gene in each cell type, combining both expression quantity and rank among genes (*82* ). Resultant gene-level expression specificity scores were then regressed with the gene-level effect sizes for imaging and psychiatric phenotypes using MAGMA gene-covariate analysis.

To map MAGMA effect sizes to snRNA-seq, we scored the single-cell enrichment by expression level of the top 1000 significant genes identified by MAGMA using single cell disease relevance score (scDRS) (*83* ). Genes were weighted by their MAGMA Z-scores, and otherwise default settings were used for scDRS analysis (using 200 bins to stratify genes by mean and variance, using estimated technical variance in expression to weight genes in computing raw score). Sensitivity analysis using top 200, 500, 2000 genes and all genes passing the FDR *<* 0.05 threshold confirmed that the scDRS single-cell enrichment scores were robust to the choice of number of significant genes to include (r *>* 0.8). Cell-level enrichment scores were then aggregated using scDRS cell-type analysis. For foetal excitatory lineage progenitors and neurons in the Wang dataset (*33* ), we further correlated the scDRS cell-level scores with the developmental pseudotime provided along with the snRNA-seq data.

### cNMF analyses

To identify gene expression programmes, we used three snRNA-seq datasets and conducted consensus non-negative matrix factorisation (cNMF) (*35* ). These datasets include the aforementioned Wang (2025) dataset (*33* ), as well as two other datasets: Braun (2023) (*36* ), which contained 111 cortical and subcortical samples from 26 individuals spanning PCW5-14, totalling 1,665,937 nuclei; Polioudakis (2019) (*84* ), which contained 4 neocortical samples from 4 individuals during PCW15-16, totalling 33,976 nuclei.

We used default parameters when running cNMF (2000 highly variable genes, 100 iterations, no regularisation) for the Wang and Braun datasets. Because the Polioudakis captures a much narrower developmental window, and is used as a sensitivity analysis for cNMF programmes in the Wang dataset, we used the same 2000 genes identified as highly variable in the Wang dataset, excluding genes with all-zero expression. We otherwise used default parameters. To identify the optimal number of components to extract, we tested the range from 5 to 40 components. In accordance with the guidelines suggested by the original cNMF publication (*35* ), we selected the optimum number of factors to maximise stability, have inter-factor correlation *<* 0.7, and to exceed the optimum number of principal components from the same dataset (elbow point), the latter of which was identified by visually examining the scree plot. Hence, we identified 16 factors for the Wang dataset, 12 factors for the Polioudakis dataset, and 18 factors for the Braun dataset. Sensitivity analyses using different numbers of factors confirmed that these factors were robust to parameter choice (**Supplementary Note**). Enrichment in each factor was then estimated using MAGMA gene-covariate regression analysis (*34* ).

To annotate the gene expression programmes to biological pathways, we obtained the non-negative gene weights on each cNMF factor, and tested the top 200 genes using enrichr (*85* ) in three gene ontology databases (biological pathway, cellular component, molecular function) (*86* ). Sensitivity analysis using 50, 100, 300 and 500 genes gave similar results. Cell-type specificity of gene expression programmes was assessed using the feature-covariate association pipeline developed in the sciRED toolbox (*37* ). The sciRED tool trains an ensemble classifier (logistic regression, decision tree and XGBoost) on the expression level of all programmes in each cell to match given cell types. Significant associations between factors and cell types were identified by automatically thresholding the feature importance scores from the classifier. Because we identified differentiation of IPCs as a putative pathway, we additionally tested the enrichment of cNMF factors in radial-glia-like versus neuronal-like IPCs using marker genes identified by ref. (*36* ). We used a permutation-based t-test over cNMF weights (100,000 permutations), where we randomly sampled the same number of genes across the 2,000 genes used to generate the cNMF weights to test this enrichment.

To assess the generalisability of cNMF programmes across datasets, particularly Wang F3, Polioudakis F3 and Braun F7, we conducted two sensitivity analyses: We first re-fit cNMF from scratch using the same set of genes across datasets, to estimate the correlation of cNMF factors identified from different datasets. We fixed the cNMF weights from one dataset and re-fit the expression activity of each factor in another dataset, which was used for cell-type classification using the sciRED toolbox (*37* ). Details of the sensitivity analyses are given in the **Supplementary Note**.

### eRegulon analyses

We tested whether ADHD and cortical expansion GWAS was enriched in computationally predicted downstream targets of the transcription factors (eRegulons) present in the cNMF factors identified in Braun (2023) and Wang (2025) datasets. To do this, we obtained eRegulons identified from multimodal snATAC-seq and snRNA-seq data (*33* ) in the developing brain using SCENIC+ (*87* ), and took the subset of transcription factors overlapping with the cNMF programmes. A 10-kb window up- and down-stream was used when mapping genes to variants. We used MAGMA (*34* ) to test for single-phenotype enrichment in each eRegulon, stratified by whether the TF had repressed or activated the target genes in the eRegulon, and used GNOVA (*38* ) to estimate the genetic covariance between psychiatric disorders and the cortical size factor within each eRegulon. The resultant genetic covariance was then correlated with cNMF weights and tested by permuting the cNMF weights, in order to identify enrichment of negative genetic overlap between cortical size and psychiatric disorders in cNMF programmes. Where one TF had both activation and repression eRegulons, we used two methods to estimate this correlation: (a) taking the sum of genetic covariance (primary method); (b) taking one eRegulon with highest absolute genetic covariance (sensitivity analyses).

### Enrichment analyses in differentiating versus proliferating progenitors

To test whether each phenotype was enriched in neural progenitors committed to differentiation or amplifying the progenitor pool, we focused on neural progenitors in the S and G2M phases of the cell cycle, when cell fate markers emerge. We determined the cell cycle phase using mean log-normalised expression of marker genes from ref. (*88* ). For cells in the S and G2M phases, we used cell-type marker genes for radial glia, excitatory IPC and newborn excitatory neurons provided by ref. (*33* ) to assign transient states. Each cell is considered positive for a cell type when the mean log-normalised expression of the corresponding cell-type marker genes exceeded 1.0.

For example, a cell is considered IPC+ Neuron+ when its mean expression of both IPC markers and neuron markers were *>* 1.0, as suggested by ref. (*84* ). A cell positive for both progenitor and neuron cell types was classified as differentiating, whereas a cell positive for only radial glia or only IPCs was classified as amplifying.

A permutation t-test was used to test differences in scDRS scores and cNMF programme activity between amplifying and differentiating progenitors, stratifying by cell cycle phase and progenitor cell type (radial glia or IPCs). Sensitivity analyses using relaxed log-normalised expression thresholds for transient states (0.1, 0.2, 0.5, 0.7) showed similar results.

### Spatial enrichment analyses

We tested the spatial specificity of cell-type enrichments by mapping the GWAS summary statistics to human mid-gestation frontal cortex spatial transcriptomic slides (**Table S24**) with gsMap (*39* ) (v1.73.6). N = 4 Visium slides representing sections from 1 individual at PCW12 and three at PCW15 included expert annotations of transient cortical and subcortical regions, which we leveraged to calculate regional enrichment for ADHD and cortical size GWAS with gsMap. Spatially informed linkage disequilibrium scores were first estimated for each spot with default settings were used (51 nearest neighbours based on gene expression, 201 spatial neighbours, and 50kb window to estimate LD score) in gsMap “quick mode”. We subsequently aggregated by brain regional annotation in the gsMap Cauchy combination test. Enrichment in brain regions was considered significant at Benjamini-Hochberg *q <* 0.05, calculated per trait-slide combination. Annotation groups with less than 30 spots in a particular slide were explicitly dropped by the gsMap Cauchy combination test.

### Mapping cNMF programmes to mid-foetal spatial transcriptomic data

We repurposed cell2location (*89* ), a popular algorithm to deconvolve cell type proportions in spot-based spatial transcriptomics such as Visium, to deconvolve cNMF programmes instead.

First, we calculated mean gene counts per cNMF programme (“cNMF signatures”) for both Wang and Braun datasets. Instead of default cell2location negative binomial regression, we calculated mean gene counts per cNMF programme as a gene-factor matrix *S*_*M,K*_: (1) weighting the snRNA-seq cell-gene count matrix *X*_*N,M*_ per cell *n* by the cNMF usage matrix (*U*_*N,K*_) per factor *k*, (2) sum across cells, (3) normalise by the total programme usage across all cells:

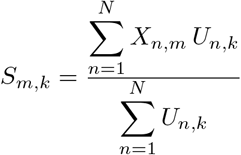

Second, cNMF signatures were used as input to cell2location (v0.1.4) deconvolution of Visium spots. Models were trained for Wang and Braun cNMF programmes per slide using default hyperparameters (`N_cells_per_location = 30, detection_alpha = 20`, 30,000 training epochs). Cell2location posterior mean estimates of cNMF programme abundance per spot were extracted, and normalised within spots to calculate per spot cNMF programme proportions.

### Human-primate DEG analyses

A spatial spin-permutation test (*74, 75* ) was used to estimate spatial correlation between ADHD-IDP genetic correlations and evolutionary cortical expansion in humans compared to macaque cortex (*90* ), similar to the colocalisation analysis with ENIGMA case-control maps as described above. To assess the specificity of our findings, we also ran spin-permutation tests with three other maps: sensory-association axis; allometric scaling, and the first principal component of gene expression the adult brain (*91, 92* ). We conducted this primarily for ADHD-SA genetic correlation, and assessed consistency in effects for ADHD-Volume/FI/and ICI genetic correlations across the cortex.

To assess if there was an enrichment in differentially expressed genes (DEGs) in snRNA-seq data from developing brains of the human and macaque cortex, or between human and chimpanzee and human cortical organoids, we used genes identified in the largest such study (*43* ). We then used MAGMA (*34* ) to estimate single-phenotype enrichment and GNOVA (*38* ) to estimate the genetic covariance between cortical size and ADHD stratified by each DEG gene set, similar to the eRegulon analysis as described above. A permutation-based t-test over cNMF weights (100,000 permutations) was used to test for enrichment in DEGs between humans and non-human primates, where we compared the mean weights of DEGs against other genes. Because multiple cNMF programmes were enriched in DEGs, we reasoned that part of these enrichments were driven by DEGs that loaded highly across multiple factors. Hence, we tested for factor-specific enrichment in DEGs by normalising the weight of each gene across factors (weight per factor/mean weight across all factors), and repeating the permutation t-test.

## Ethics statement

All ethical procedures for UK Biobank (UKB) are controlled by the UKB Ethics Advisory Committee. Additionally, the current study followed the UKB Ethics and Governance Framework document (https://www.ukbiobank.ac.uk/ethics-and-governance-framework/), with approval from the Northwest Multi-centre Research Ethics Committee (MREC), as a Research Tissue Bank (RTB) approval. This approval means that researchers do not require separate ethical approval for accessing and analysing de-identified data as was done in this study, and can operate under the RTB approval.

We ensured compliance with the terms and conditions outlined for accessing summary statistics from the Psychiatric Genomics Consortium (PGC). These terms and conditions can be found on the PGC website at the following link: https://pgc.unc.edu/for-researchers/download-results/

## Acknowledgements

We would like to thank Daniel Geschwind (University of California, Los Angeles) for his constructive advice on this research.

OAB was funded by Wellcome Sanger core funding (220540/Z/20/A) and a grant from SFARI (736282). VW, NW and ETB are supported by ImmunoMIND (MR/Z50354X/1), a hub in the national UKRI Mental Health Platform. VW, DA, M-E L, SA, SR, MG, and ETB are supported by ANXIOMICS, a grant from the Wellcome Trust (309245/Z/24/Z). VW and SBC receive funding from the European Unions Horizon 2022 R2D2-Mental Health project, SFARI, and the Wellcome Trust (214322/Z/18/Z). All research at the Department of Psychiatry in the University of Cambridge is supported by the NIHR Cambridge Biomedical Research Centre (BRC-1215-20014) and NIHR Applied Research Centre. The views expressed are those of the author(s) and not necessarily those of the NIHR or the Department of Health and Social Care. This work was supported by the John Lambton Trust fund and by the K. Lisa Yang Centre for Autism Research at Cambridge. Any views expressed are those of the author(s) and not necessarily those of the funders. The funders had no role in the design of the study, in the collection, analyses, or interpretation of data, in the writing of the manuscript, or in the decision to publish the results. For the purpose of Open Access, the authors have applied a CC BY public copyright licence to any Author Accepted Manuscript version arising from this submission.

## Author contributions

YH, AE and VW conceptualised and designed the study; YH and YG quality-controlled the genotype data; AE, YG, RR-G and RAIB pre-processed and quality-controlled the imaging data; YG conducted the GWAS; AE, CMLR and BP investigated genetic correlation and PGS; YH and VW performed MR, cell-type, programme and evolutionary enrichment analyses; KR, EJ, FF and OAB generated spatial transcriptomic data and conducted spatial enrichment analyses; YH and AE performed statistical analyses (matrix decomposition and permutation tests); RP, JG, ADB, NP, OA, OF, DC, NB provided data for this study; AL, JZ and NW provided software for this study; SB-C, MG, SA, M-EL, DA, RAIB, ETB, OAB and VW acquired funding; MG, SA, SR, DA and VW were involved in patient and public involvement and engagement (PPIE) of this study; TR, RAIB, OAB and VW supervised this study; YH, AE and VW wrote the first draft, generated the figures and proofread the final submission; all authors provided feedback for the article.

## Data availability

Raw imaging and genetic data can be obtained from UK Biobank (www.ukbiobank.ac.uk), ABCD (abcd-study.org) and dHCP (www.developingconnectome.org). Summary statistics for the cortical size common factor will be made available upon publication.

## Code availability

Standard pipelines for genetic correlation, polygenic scoring, matrix decomposition, Genomic SEM, MAGMA enrichment, Mendelian randomisation, and spatial co-localisation are available at:

- https://github.com/amir-ebneabbasi/Brain-dynamics-GWAS (Genetic correlation and polygenic scoring)
- https://github.com/amir-ebneabbasi/Matrix-decomposition (Matrix decomposition)
- https://github.com/yh464/gwas_pipeline (Genomic SEM, enrichment and Mendelian randomisation)
- https://github.com/amir-ebneabbasi/Spatial-Colocation (Spatial co-localisation)

Pipelines for multiomic data preprocessing, Cepo scoring, and cNMF decomposition are available at:

- https://github.com/yh464/multiomics_pipeline

Ad-hoc analyses and plotting scripts used in this study are available at:

- https://github.com/yh464/2025-img-func-genomics

Quality control pipeline of dHCP genotyping data is available at:

- https://github.com/yh464/dhcp_genotype_qc

