## Supplementary Figures and Notes for "Polygenic effects on cortical size and ADHD converge on a mid-gestational progenitor-to-neuron transition programme"

[1]Department of Psychiatry, University of Cambridge, Cambridge, UK
[2]Department of Clinical Neurosciences, University of Cambridge, Cambridge, UK
[3]Wellcome Sanger Institute, Hinxton, UK
[4]Institute of Psychiatry, Psychology and Neuroscience, King's College London, London, UK
[5]Department of Metabolism, Digestion and Reproduction, Imperial College, London, UK
[6]Department of Psychiatry, Yale University, New Haven, USA
[7]The Lundbeck Foundation Initiative for Integrative Psychiatric Research, iPSYCH, Aarhus, Denmark
[8]Center for Genomics and Personalized Medicine (CGPM), Aarhus University, Aarhus, Denmark
[9]Department of Biomedicine (Human Genetics) and iSEQ Center, Aarhus University, Aarhus, Denmark
[10]Bioinformatics Research Centre, Aarhus University, Aarhus, Denmark
[11]Centre for Precision Psychiatry, Division of Mental Health and Addiction, University of Oslo and Oslo University Hospital, Oslo, Norway
[12]Division of Psychological Medicine and Clinical Neurosciences, University of Cardiff, Cardiff, UK
[13]Department of Psychiatry, University of Oxford, Oxford, UK
[14]Department of Medical Physiology and Biophysics, Institute of Biomedicine of Seville (IBiS) HUVR/CSIC/University of Seville/CIBERSAM, ISCIII, Seville, Spain.
[15]Department of Women's and Children's Health, University of Liverpool, Liverpool, UK
[16]Alder Hey Children's NHS Foundation Trust, Liverpool, UK
[17]Cambridge Mind and Brain Institute, University of Cambridge, Cambridge, UK
[18]Department of Psychology, University of Cambridge, Cambridge, UK
[19]UCL Cancer Institute, University College London, London, UK
[**]These authors contributed equally: Y.H, A.E.

### Supplementary Figures

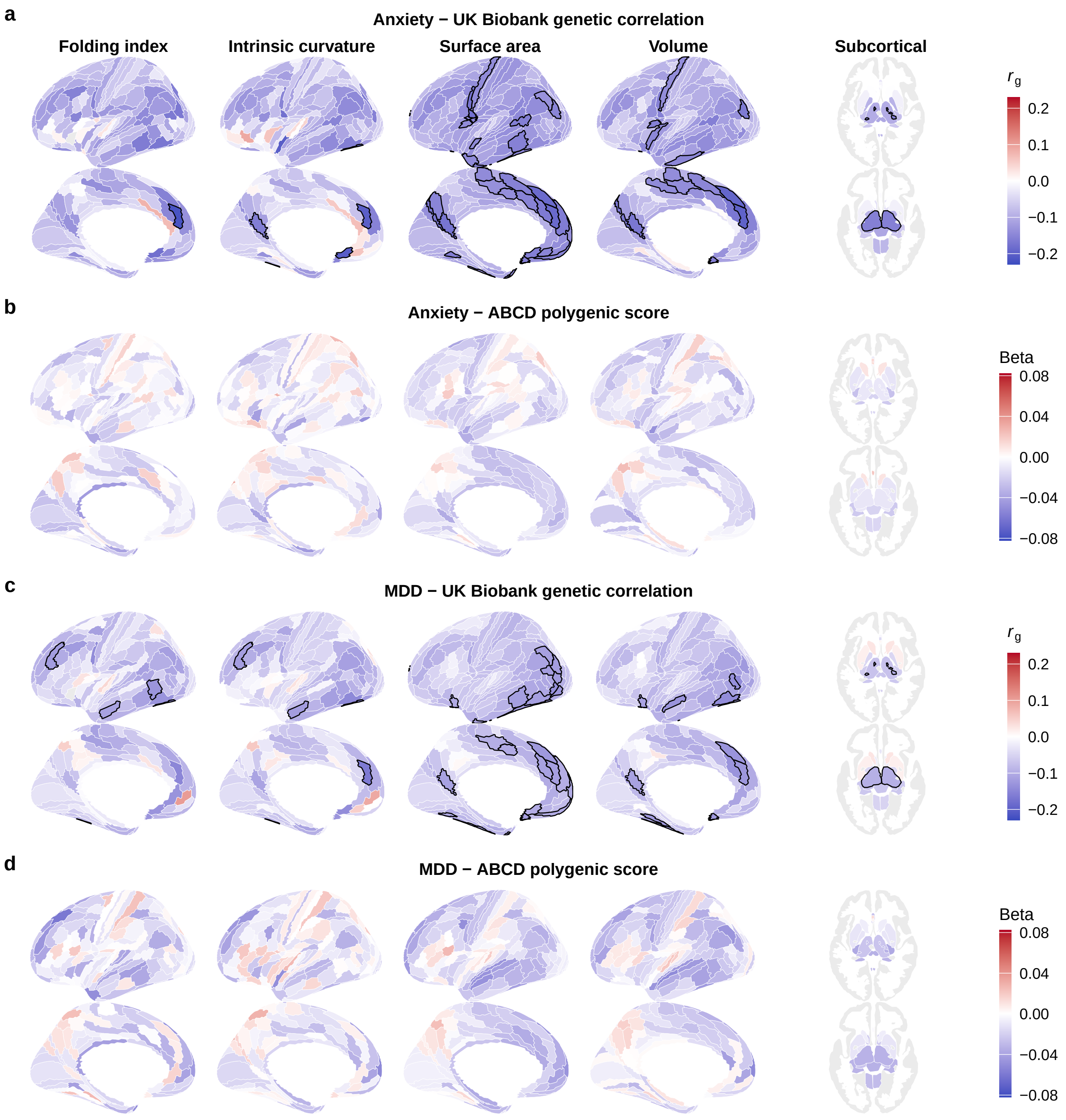


Figure S1: **Genetic overlap between depression, anxiety and imaging phenotypes.** **a** Genetic correlation $r_g$ with anxiety, estimated using imaging GWAS based on the UK Biobank. **b** Anxiety polygenic score effect size $\beta$ on imaging phenotypes for individuals in ABCD. **c-d** Same as **a-b** but for major depression (MDD).

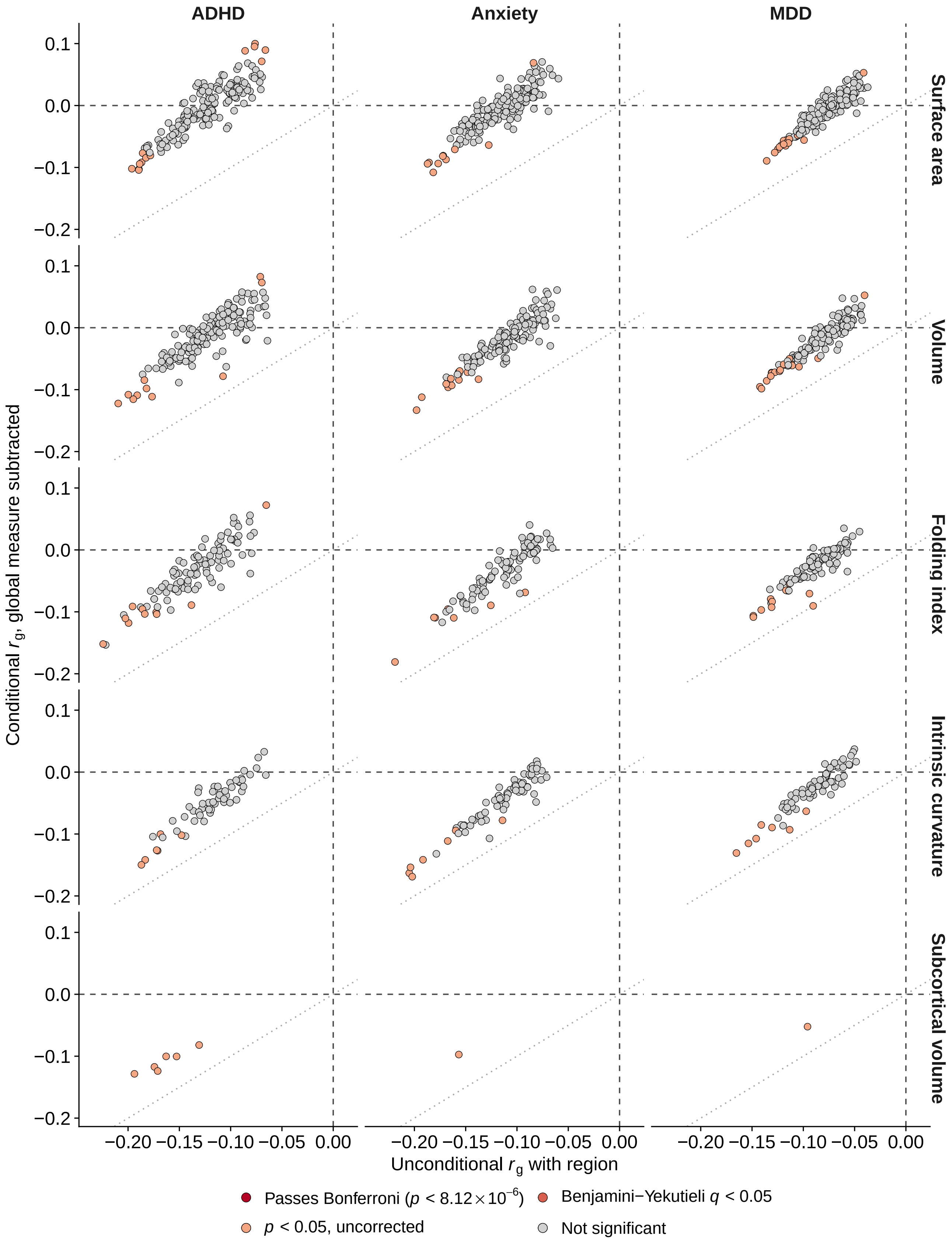


Figure S2: **Genetic correlation $r_g$ between regional IDPs and psychiatric disorders,** adjusted versus unadjusted for the corresponding global IDP, based on imaging GWAS within the UK Biobank. The dashed lines are identity lines. Regions are colour coded by the significance of the adjusted $r_g$.

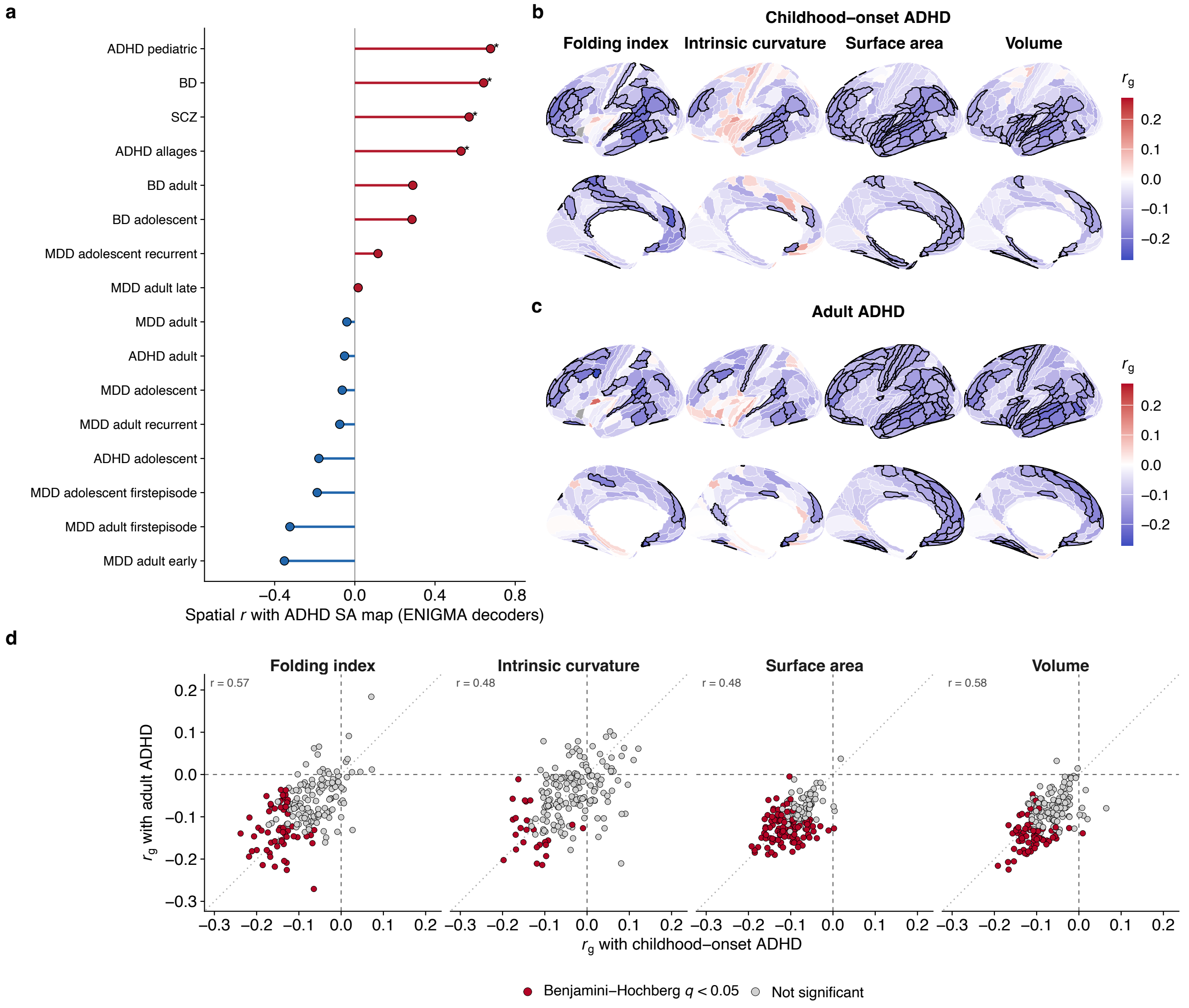


Figure S3: **Sensitivity analysis for genetic correlation between cortical size and ADHD. a** Spatial co-localisation between ADHD polygenic score effect sizes on cortical surface area and ENIGMA case-control difference maps in cortical surface area. BD: bipolar disorder; SCZ: schizophrenia; MDD: major depressive disorder. *: passing the Benjamini-Hochberg $q < 0.05$ threshold. **b-c** Genetic correlation $r_g$ between regional IDPs and (**b**) childhood and (**c**) adult-onset ADHD. Dark borders indicate regions passing the $p < 8.12 \times 10^{-6}$ Bonferroni threshold. **d** Genetic correlation $r_g$ between regional IDPs and childhood- versus adult-onset ADHD. Each phenotype is colour coded by the significance of its genetic correlation with *all* ADHD.

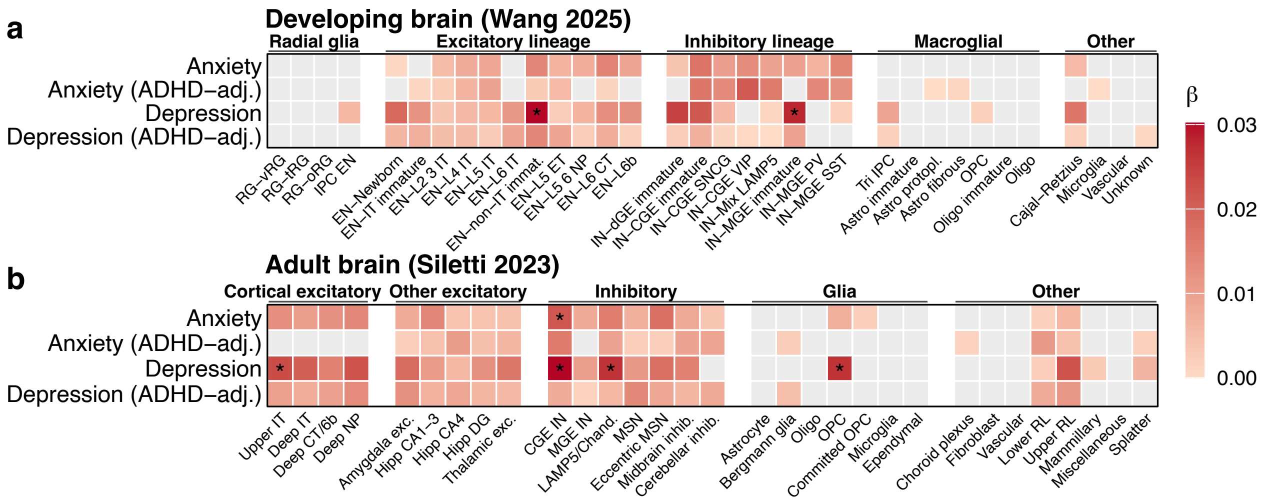


Figure S4: **Cell-type enrichment of anxiety and depression**, unconditional and adjusted for ADHD, in the (**a**) developing and (**b**) adult brain. Enrichment is estimated by correlating GWAS effect sizes with gene expression specificity scores for each cell type derived using Cepo. *: passing the Benjamini-Hochberg $q < 0.05$ threshold. Non-significant ($p > 0.05$) associations are coloured grey.

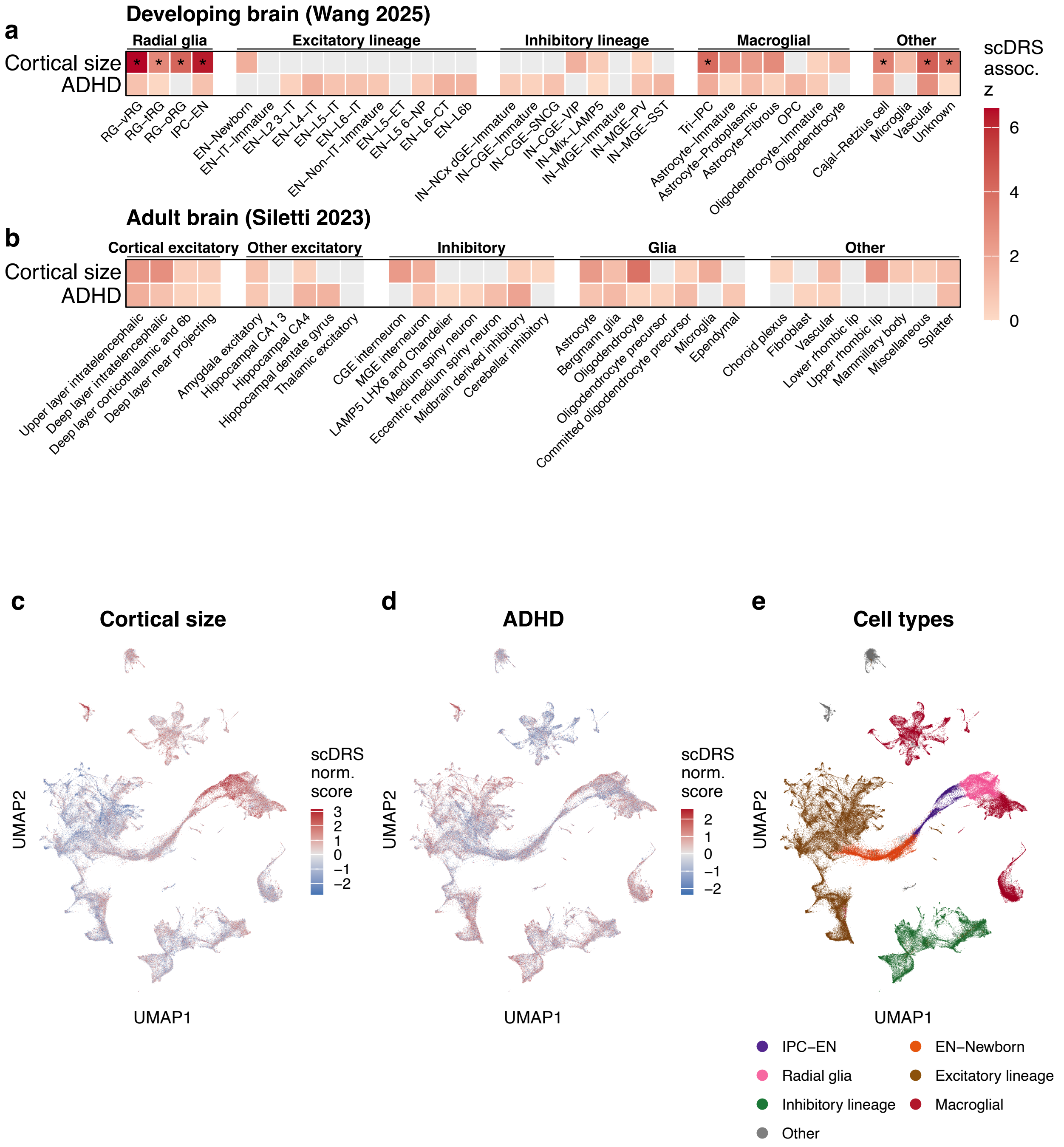


Figure S5: **Enrichment of ADHD and cortical size estimated using scDRS. a-b** Cell-type enrichment in the (**a**) developing and (**b**) adult brain. *: passing the Benjamini-Hochberg $q < 0.05$ threshold. Non-significant ($p > 0.05$) associations are coloured grey. **c-d** Cell-level enrichment score in the developing brain for (**c**) cortical size and (**d**) ADHD, as estimated using single-cell disease enrichment score (scDRS), plotted in UMAP space. **e** Cell-type legend re-drawn using data from Wang et al (2025). IPC: intermediate progenitor cell; EN: excitatory neuron.

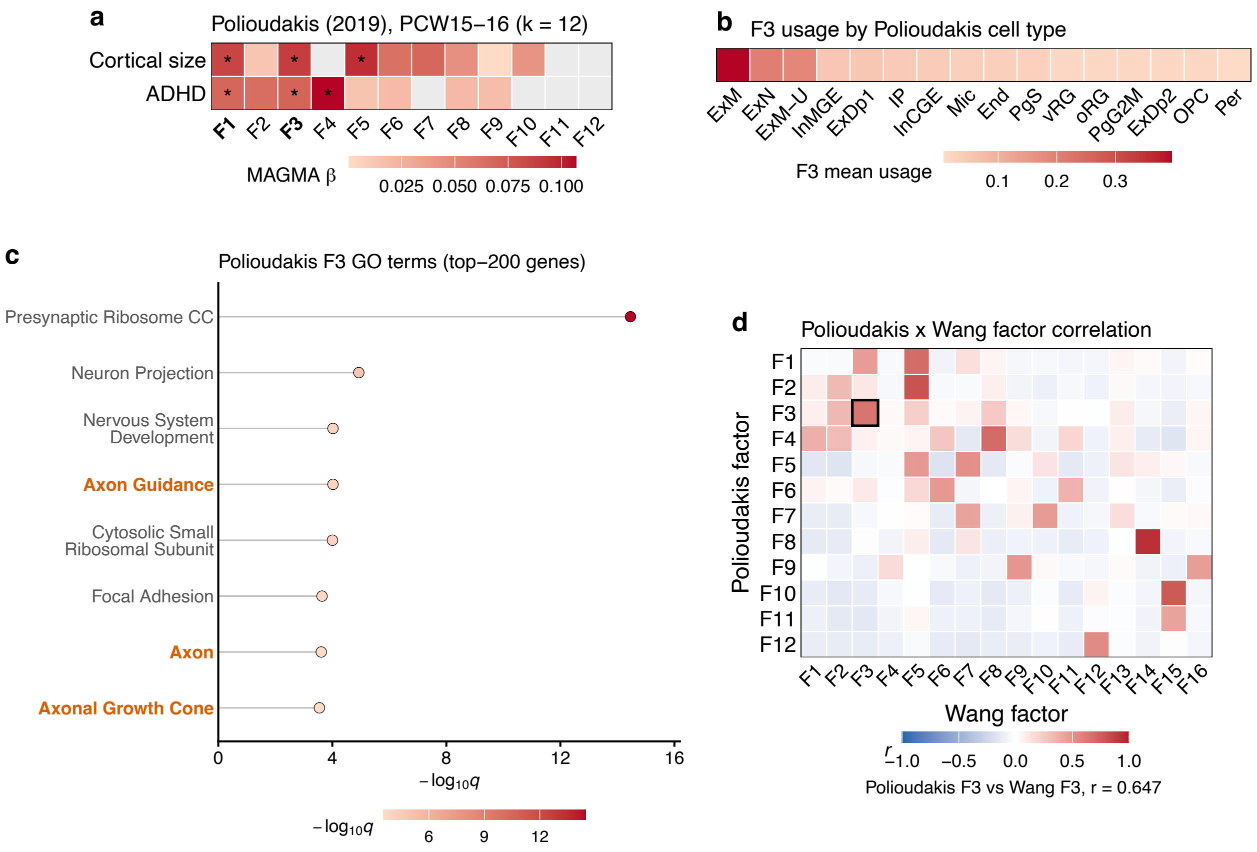


Figure S6: **Replication of Wang F3 in Polioudakis midgestation dataset.** **a** MAGMA-based enrichment of ADHD and cortical size GWAS in the 12 cNMF factors. *: passing the Benjamini-Hochberg $q < 0.05$ threshold. **b** Mean cell-level activity of Polioudakis F3 factor by cell type, in decreasing order. ExM: migrating excitatory neurons; ExN: excitatory neurons. **c** Gene ontology enrichment for the top 200 genes of Polioudakis F3 factor. **d** Correlation between cNMF factors of the Polioudakis dataset and cNMF factors of the Wang dataset.

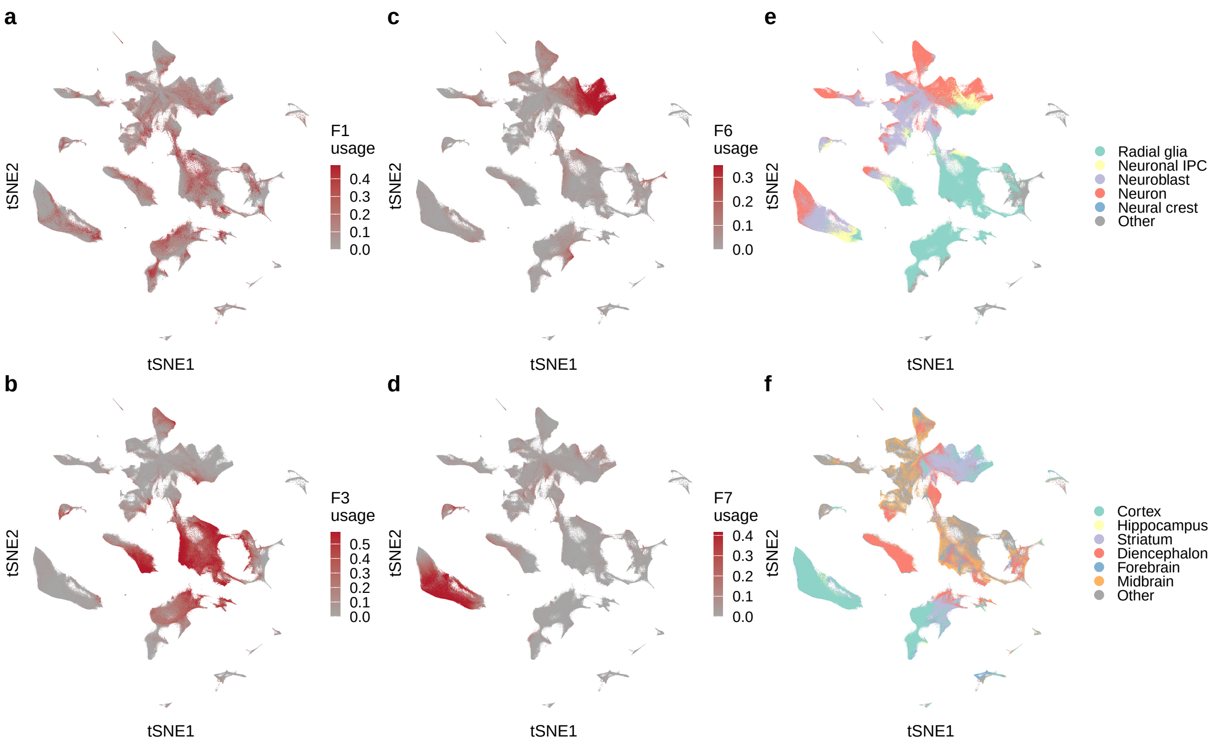


Figure S7: **Expression activity of Braun cNMF factors in tSNE space,** for the four overlapping factors between ADHD and cortical size: (**a**) F1, (**b**) F3, (**c**) F6 and (**d**) F7. Legends for (**e**) cell types and (**f**) brain regions are re-drawn using original data from Braun et al (2023) (*1*).

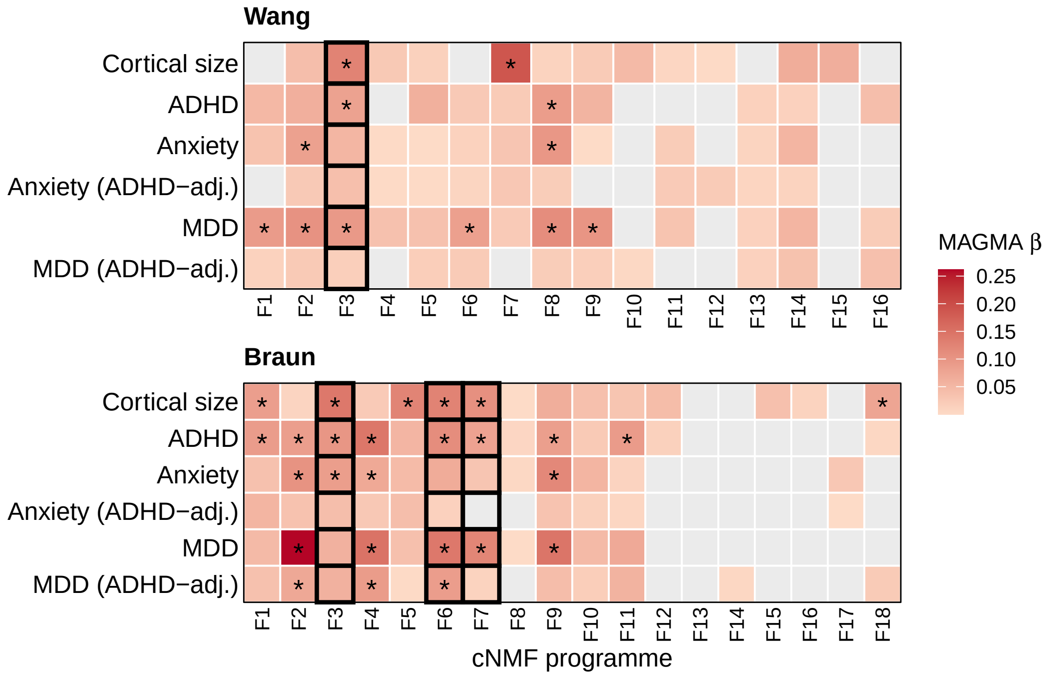


Figure S8: **Enrichment of depression and anxiety in Wang and Braun cNMF programmes,** unadjusted and adjusted for ADHD, as estimated using MAGMA. *: passing the Benjamini-Hochberg $q < 0.05$ threshold. Non-significant (nominal $p > 0.05$) enrichments are coloured grey. Enrichment for ADHD and cortical size are provided for comparison. The four key programmes (Wang_F3, Braun_F3, Braun_F6, Braun_F7) overlapping between cortical size and ADHD are shown in bold.

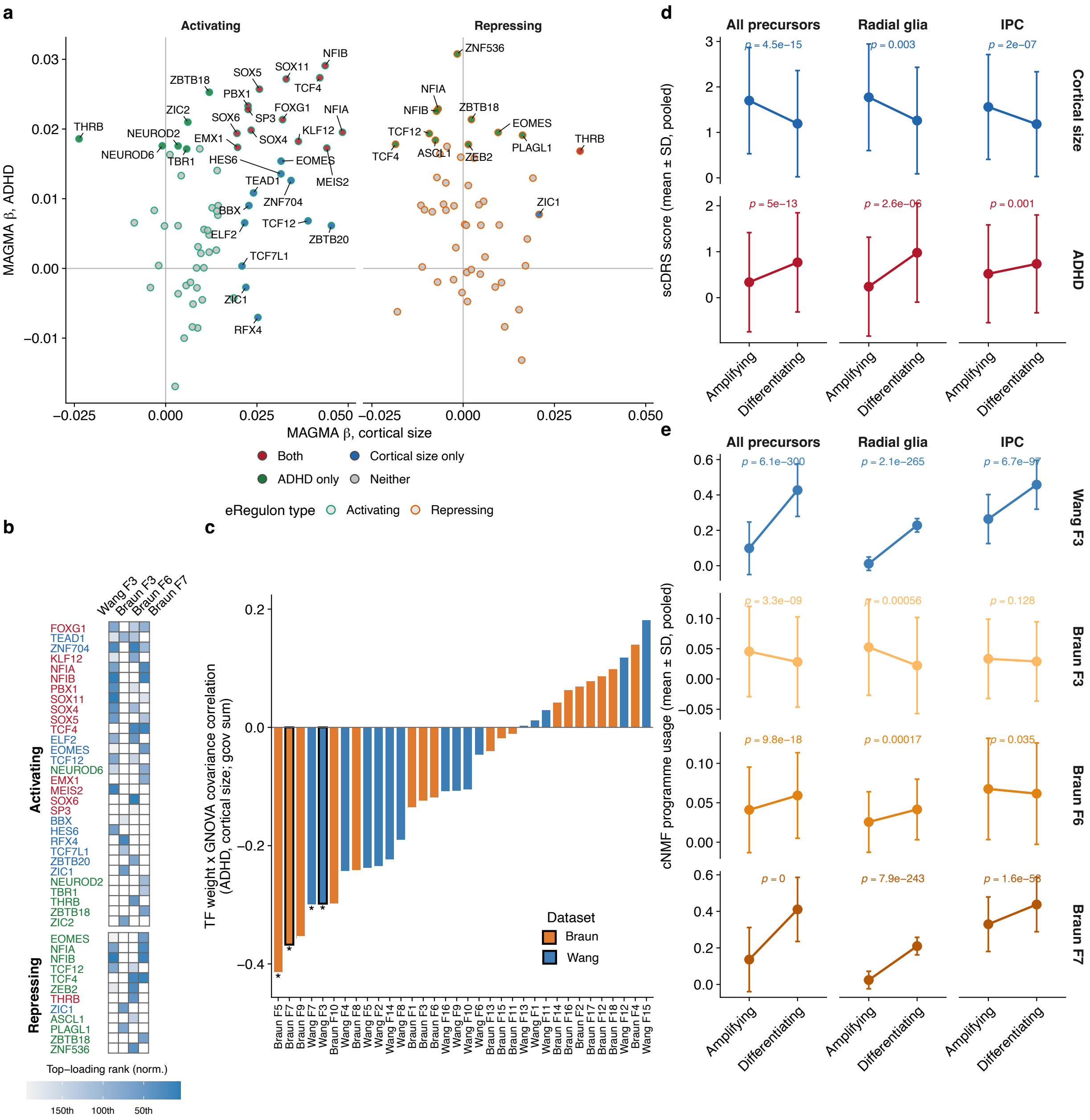

Figure S9: **Enrichment in downstream targets of transcription factors (eRegulons).** **a** Single-trait enrichment for ADHD and cortical size in each eRegulon estimated using MAGMA, colour-coded by significance at the Benjamini-Hochberg $q < 0.05$ threshold. Activating and repressing targets of transcription factors are plotted separately. **b** Overlap between top significant eRegulons and the top 200 genes of cNMF factors (Wang F3, Braun F3, F6, F7). Genes not in the top 200 are left blank. **c** Correlation between cNMF weights of each transcription factor and the genetic covariance (ADHD with cortical size) within its downstream eRegulon estimated using GNOVA. Significance is determined by permuting the cNMF weights. *: passing the Benjamini-Hochberg $q < 0.05$ threshold. **d** scDRS scores of ADHD and cortical size in amplifying versus differentiating neural progenitors, mean ± standard deviation. **e** cNMF factor expression activity in amplifying versus differentiating neural progenitors, mean ± standard deviation.

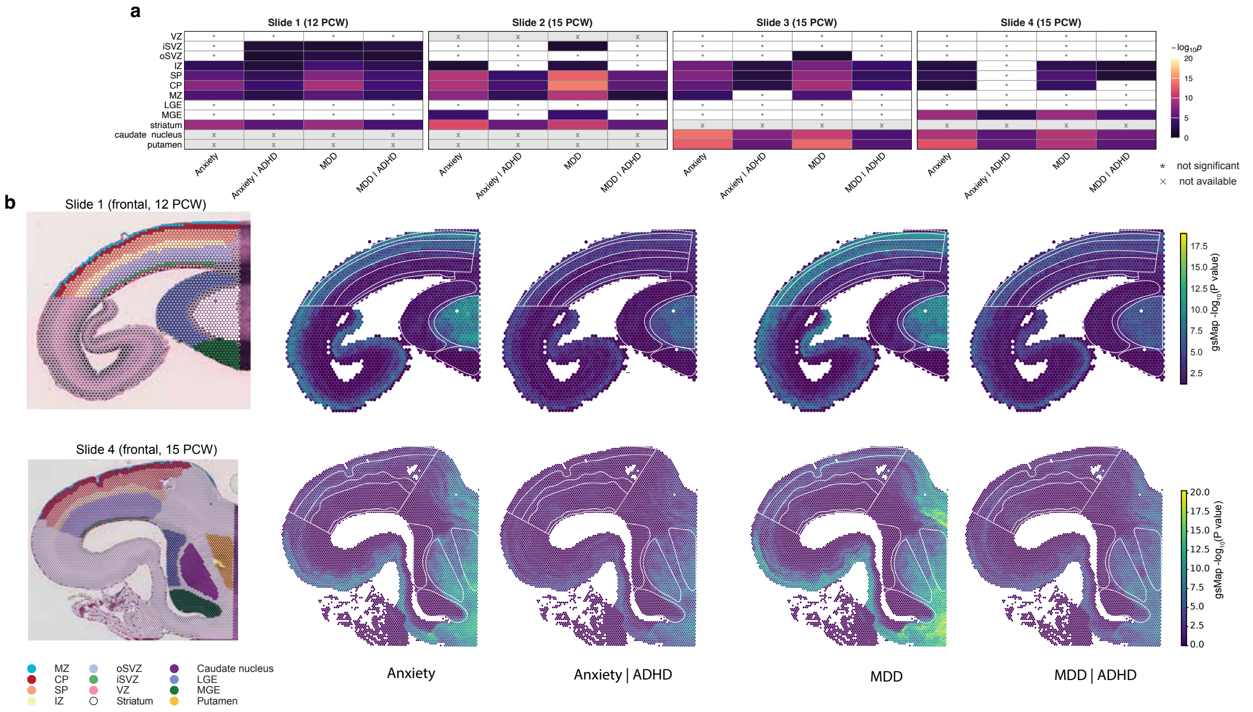


Figure S10: **Anxiety and MDD show limited, inconsistent enrichment in cortical proliferative zones, unlike ADHD and cortical size.** **a** Regional gsMap enrichment for anxiety and MDD, raw and ADHD-adjusted, across three spatial transcriptomic sections (GW14: AADOW 13042003; GW17: AADOW 13458215, AADOW SP13163480). Spot-level p-values were combined per annotated zone via the Cauchy combination test; tile colour is -log10(p) (capped at 20, same scale as Figure 6). Grey: tested, not significant (FDR $q$ <0.05 within each section × trait); blank: region not annotated in that section. Rows are ordered developmentally, cortical zones (VZ, iSVZ, oSVZ, IZ, SP, CP, MZ) then subcortical/ganglionic structures (LGE, MGE, striatum, caudate, putamen), marked by the left-hand colour bar. **b** The underlying gsMap spot-level maps for the same four traits and sections, with each section's region-annotation index for reference. Colour scales represent within-slide enrichment p-values and are not comparable across phenotypes or across slides. Neither anxiety nor MDD showed consistent enrichment in proliferative zones, unlike ADHD and cortical size (Figure 6a-b) both were instead concentrated in postmitotic/subcortical regions - indicating specificity, not just lower GWAS power, drives ADHD's proliferative-zone signal.

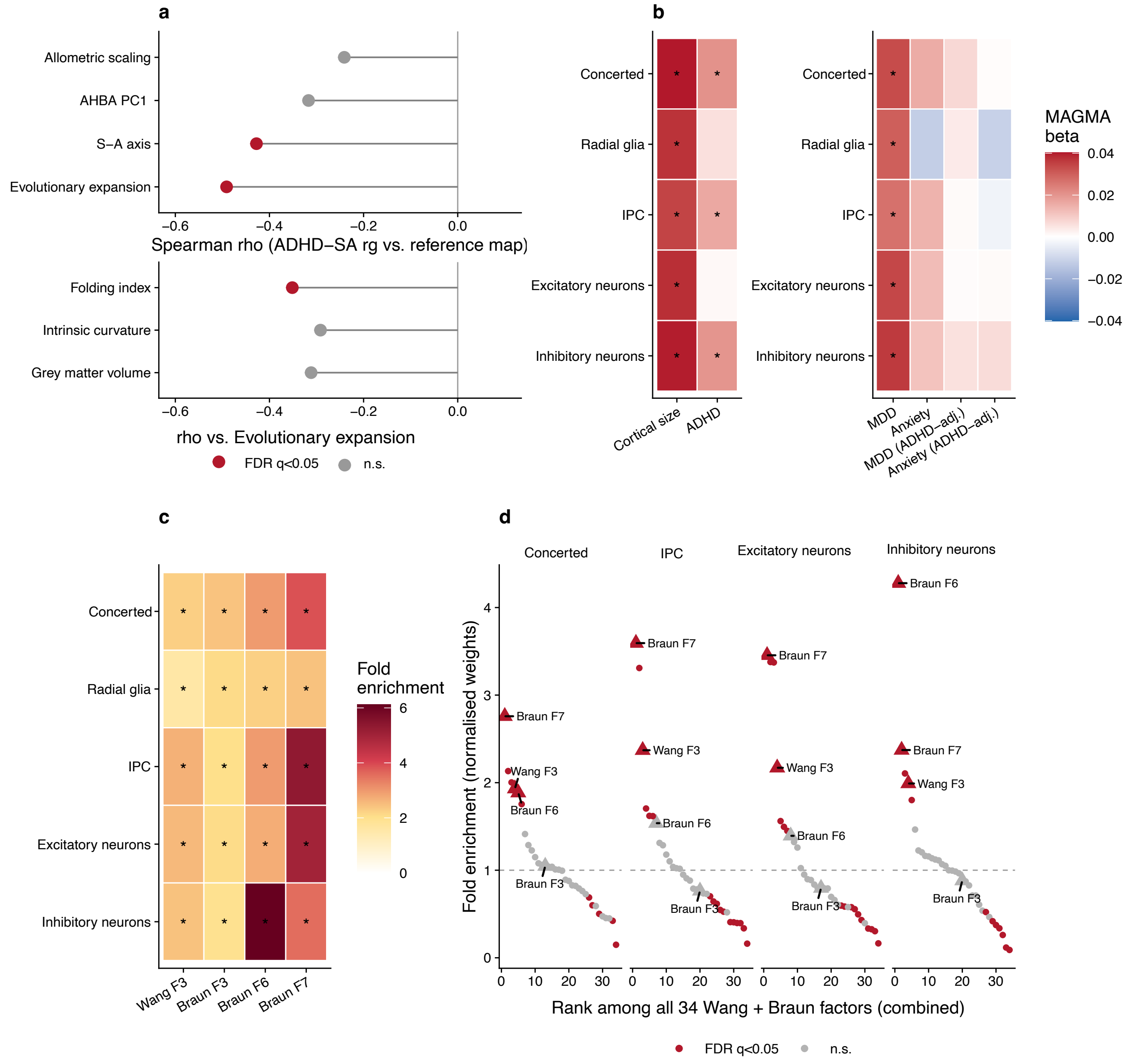


Figure S11: **Enrichment of the genetic overlap between ADHD and cortical size in evolutionary signatures.** **a** Spatial correlation between the regional surface area-ADHD genetic correlation map and evolutionary cortical expansion of human relative to macaques, compared with control topographic brain maps (top) and genetic correlation maps of other IDPs with ADHD (bottom). Significant spatial correlations are coloured red. **b** Enrichment of GWAS of cortical size, ADHD, depression and anxiety (unadjusted and adjusted for ADHD) in differentially expressed genes (DEGs) between human and macaques, estimated using MAGMA. **c-d** Enrichment of the cNMF programmes in human-macaque DEGs, using a permutation t-test on (**c**) raw cNMF weights and (**d**) normalised cNMF weights (gene-level weight / mean weight across all factors for that gene). Fold enrichment = the mean weight of DEGs / mean weight of permuted gene sets. Concerted: genes that are differentially expressed across all cell types. *: passing the Benjamini-Hochberg $q < 0.05$ threshold.

### Supplementary Note: Consensus NMF preliminary and sensitivity analyses

#### Parameter selection for cNMF

For each scRNA-seq dataset, we took the subset of protein-coding genes, because downstream MAGMA gene-covariate analysis was based on protein-coding genes only. We then selected the top 2000 highly variable genes (HVGs, the default setting of cNMF) and conducted cNMF decomposition. We did an exhaustive search from K = 5 components to K = 40 components. Within that range, we selected K based on the principles recommended by the original publication of cNMF (*2*):

1. Number of components should be at least the optimal number of principal components (over the same set of HVGs), as visually determined by the PCA scree plot.

2. Number of components should represent a peak in stability (Silhouette score).

3. Components should be as uncorrelated as possible, in particular, we only selected K such that the correlation between each pair of factors $r < 0.7$.

Close decisions were made by judgement. In particular, we correlated factors produced by multiple candidate K values to identify if factors were consistent across parameter choice. We also inspected the cell-type specificity by visual inspection of the expression of each factor, and training an ensemble classifier over cell type labels as in the sciRED pipeline (*3*). A suitable K value was selected balancing resolution and biological interpretability. After K is determined, the local density threshold (DT) parameter was determined by visual inspection of the spectral histogram.

For the Wang (2025) dataset (*4*), the optimal number of principal components was 11, and we identified two peaks in stability above 11 (K = 16, 35) (**Figure S12a**). Because there were only 33 cell types with several closely related groups, we selected K = 16 for better stability and better biological interpretability. Visual inspection of the spectral histogram confirmed that 0.1 was a suitable density threshold (**Figure S12b**). We confirmed that the correlation between factors ranged from -0.159 to +0.694, within the acceptable threshold. In particular, the overlapping factor between ADHD and cortical size (F3) is only moderately correlated with other factors (r = -0.127 to +0.286, Table S16). We also tried K = 10 which had better stability, but the biological resolution was not good (one factor identifies both OPCs and mature excitatory neurons; another factor identifies differentiated neurons and radial glia but not intermediate progenitors). Hence, we proceeded with K = 16 and DT = 0.1 for further analyses. We tested a range of K values (13 to 19) and correlated the factors with K = 16 as sensitivity analyses, and found highly correlated factors for every factor in K = 16 (**Table S30a**). Hence, we are confident that the biological programmes were robust to parameter choice.

The Polioudakis (2019) dataset (*5*) was intended as a sensitivity analysis for cNMF in the Wang (2025) dataset, but because it covers a much narrower time window, only 248 out of 2000 highly variable genes (HVGs) overlapped with the HVGs of Wang (2025). Hence, we took the 1929 HVGs of Wang that were sequenced in the Polioudakis dataset and fit cNMF on that subset. For this subset, the optimal number of principal components was 8 (**Figure S12a**) and we identified two major peaks in stability (K = 9, 10, 12). These candidates had very similar ranges of factor correlation (K = 9, r = -0.149 to +0.594; K = 10, r = -0.139 to +0.595; K = 12, r = -0.133 to +0.573). Correlating across these K values confirmed the stability of these factors, but K = 12 identifies a factor not corresponding to any factor from K = 10 or K = 9 (**Table S30b**). Hence, we proceeded with K = 12 for further analyses. We confirmed that selecting a density of threshold of 0.1 was appropriate from the spectral histogram (**Figure S12b**).

For the Braun (2023) dataset (*1*), the optimal number of principal components was 9 (**Figure S12a**) and we identified one major peak in stability. K = 18 and K = 19 had very similar Silhouette scores but the factor correlation for K = 18 (r = -0.166 to +0.454, Table S16) was much lower than K = 19 (r = -0.166 to +0.633). Correlating factors from K = 18 and K = 19 showed that this increased correlation results from two factors (K19.F15 and K19.F19) mapping onto the same factor in K = 18 (F14). Other factors are highly stable across this parameter choice (**Table S30c**). Hence, we selected K = 18 for further analyses. A visual inspection of the spectral histogram suggested that 0.1 was a suitable density threshold (**Figure S12b**).

#### Robustness of Wang F3

Because the Wang dataset spanned a great diversity of cell types, in order to assess the stability of F3 using HVGs at finer biological resolution, we re-fit cNMF on the subset of neural progenitors (radial glia and IPCs),

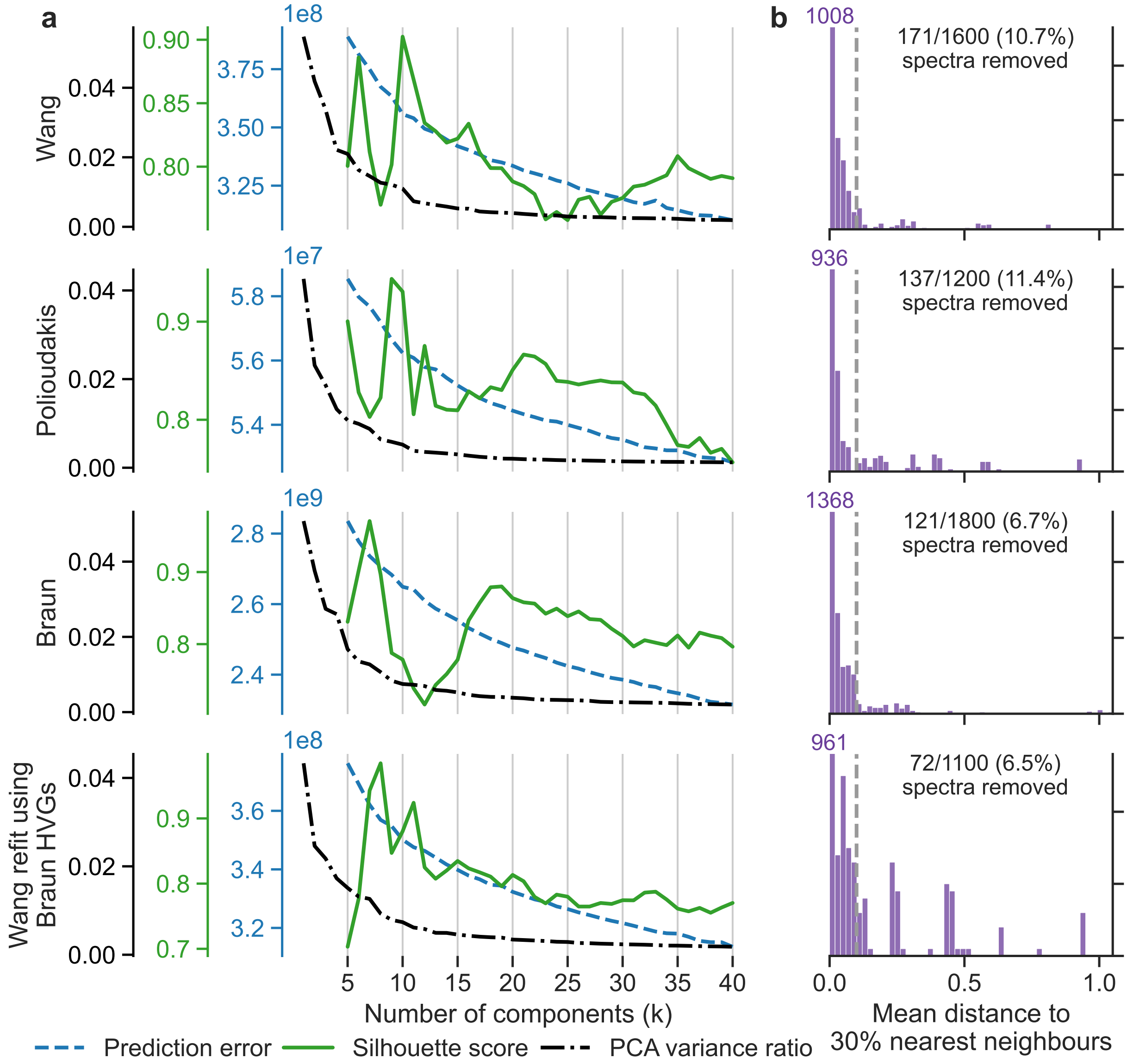


Figure S12: **Parameter selection for cNMF. a** Statistics used to select the number of components (K) for cNMF, including prediction error, stability (silhouette score) and the variance explained by the k-th principal component. **b** Local density histogram for determining the local density threshold, for K = 16 (Wang), 18 (Braun), 10 (Polioudakis) and 11 (Wang refit using Braun HVGs). Local density is defined as the mean distance to 30% nearest gene spectra. The dashed line is the selected local density threshold (0.1). The left-most bin exceeds the upper limit of the axis and the actual number of spectra in that bin is labelled.

and on the subset of F3-related cell types (IPCs and newborn excitatory neurons). For the progenitors subset, 1133/2000 HVGs overlapped with the full dataset and we identified 9 factors. One factor (rgipc_F3) was strongly correlated with the full F3 ($r = 0.835$) and nothing else ($\max |r| = 0.213$). For the IPC/newborn excitatory neurons subset, 1127/2000 HVGs overlapped and we identified 11 factors. One factor (ipcenn_F1) was strongly correlated with the full F3 ($r = 0.719$) and nothing else ($\max |r| = 0.135$). Two more factors (ipcenn_F2,F3) were correlated with both the full F3 ($r = 0.538, 0.600$) and differentiated neuron-related factors (F2, F8, $r > 0.3$, **Table S30d**).

We also re-fitted cNMF to the full Wang dataset including non-coding RNA, where 1605/2000 HVGs overlap and we found 16 factors. One factor (Wang_full_F2) was almost identical to F3 in the protein-coding genes ($r = 0.99922$, **Table S30d**). Overall, these results suggest that the co-expression of Wang_F3 is robust to the choice of HVGs.

#### Mapping between scRNA-seq datasets

Because only 923 out of 2000 HVGs overlapped between the Braun and Wang datasets, in addition to correlations between cNMF weights and overlapping gene ontology (GO) enrichments, we conducted additional analyses to assess the biological overlap between gene expression programmes identified in Wang and Braun datasets.

First, we projected the gene expression programmes identified in Braun: Out of 2000 HVGs in Braun, 1810 were sequenced in Wang. We took the raw counts of this subset and log-normalised using the same procedure as fitting cNMF from scratch. We then fixed the components matrix from Braun (taking the 1810-gene subset, $1810 \times 18$) and and fitted the normalised count matrix ($1810 \times N$) to the components matrix to get the activity of each programme in each cell ($18 \times N$). The cell-level activity of the projected programmes were subsequently analysed for cell-type specificity using an ensemble classifier (*3*), and for temporal dynamics by conducting quadratic regression against pseudotime and age, using the same methods as other from-scratch cNMF factors.

Consistent with the correlation with Wang.F3 and enrichment in early neurogenesis GO terms, projected Braun.F7 was enriched in extitatory intermediate progenitor cells (IPCs) and newborn excitatory neurons, in addition to some deep-layer neuronal subtypes (**Table S31a**). The temporal trajectory of Braun.F7 coincided with Wang.F3, peaking at around post-conception week (PCW) 14 (**Figure 5h**), and is continuous with the increasing trajectory during the first trimester in the Braun dataset (**Figure 5f**).

To assess whether this projection reflects genuine gene co-expression in the Wang dataset, we further re-fit cNMF in the Wang dataset using the same 1810 genes. We identified 11 factors at a density threshold of 0.1 (**Figure S12**) and one factor (F3) strongly correlated with Braun.F7 ($r = 0.835$, **Table S31b**), confirming the reliability of this gene co-expression programme. Braun.F3 and Braun.F6 did not correlate with any factor in the Wang dataset (using Wang HVGs, **Table S16**), nor with any factor re-fit using Braun HVGs. This likely reflects the low sampling rate of the Wang dataset in the first trimester.
